# Pulmonary fibrosis after COVID-19 is characterized by airway abnormalities and elevated club cell secretory protein-16

**DOI:** 10.1101/2025.09.08.25334667

**Authors:** Matthew R. Baldwin, Ansley E. Jones, David Zhang, Chandan Gurung, Zain Khan, Anjali Saqi, Xuehan Yang, Ying Wei, Renu Nandakumar, Scarlett O. Murphy, Claire F. McGroder, Faisal Shaikh, Selim Arcasoy, Luke Benvenuto, Harpreet Grewal, Benjamin M. Smith, Agnes CY Yuen, Parteek Johal, Chrisopher Carlsten, Christopher J Ryerson, J. Brent Richards, Alyson W Wong, Tomoko Nakanishi, Aditi S Shah, Christine Kim Garcia

## Abstract

Prior studies testing biomarkers of residual lung abnormalities after COVID-19 are limited by sampling within the first year after acute COVID-19 illness and lack of external validation of findings. In three independent, international, racially and ethnically diverse prospective cohorts of survivors of moderate to critical COVID-19, we systematically tested 18 circulating biomarkers of inflammation, aging, endothelial activation, pulmonary epithelial function, fibrosis, and fibrinolysis. We found that only higher club-cell secretory protein-16 (CC16) levels are consistently associated with persistent fibrotic lung abnormalities in cross-sectional and longitudinal analyses for up to 3 years after acute COVID-19. Histopathological and single-cell RNA sequencing analyses of transbronchial biopsies of fibrotic lung abnormalities in COVID-19 survivors sampled between 3 and 4 years after acute illness and of COVID-19 lung explants suggest that circulating CC16 reflects underlying deranged pulmonary epithelial progenitor proliferation and anomalous CC16/MUC5B-related pro-fibrotic signaling in the distal airways. CC16 should be investigated further as a potential blood biomarker that may facilitate screening of COVID-19 survivors for pulmonary fibrosis and its progression.

## INTRODUCTION

Some survivors of coronavirus disease 2019 (COVID-19) have residual abnormalities consisting of persistent reticulations and traction bronchiectasis that are similar to radiographic findings of idiopathic pulmonary fibrosis (IPF).^1^ Genome-wide association studies similarly suggest a shared genetic etiology between COVID-19 and IPF.^2,3^ In COVID-19 ARDS, there is an accumulation of profibrotic macrophage populations that are also found in IPF,^4^ and a time-dependent shift from pro-inflammatory to pro-fibrotic gene expression patterns.^5,6^

Several longitudinal cohort studies of adults hospitalized with COVID-19 suggest that 30-75% have persistent radiographic patterns consisting predominantly of reticulations and traction bronchiectasis that persist during the first year after acute SARS-CoV-2 infection.^1,7–13^ Greater initial severity of illness and receipt of invasive mechanical ventilation are major clinical risk factors for fibrotic-like abnormalities after COVID-19.^7–9,14,15^ Reassuringly, fibrotic-like abnormalities are usually associated with only mild reductions in diffusion capacity, and they are not consistently associated with restrictive or obstructive ventilatory defects, reduced exercise tolerance, or dyspnea.^7,9,10,13,14^ However, these findings may be falsely reassuring since pulmonary fibrosis is usually an insidious sub-clinical process that becomes symptomatic only after lung function has declined substantially.^16^ To date, there are no known post-acute blood-based biomarkers of post-COVID fibrotic lung patterns that offer mechanistic insight or prognostic enrichment for these concerning thoracic imaging findings of fibrosis.

Biomarkers of inflammation, aging, endothelial activation, pulmonary epithelial function, fibrosis, and fibrinolysis are associated with COVID-19 acute lung injury (ALI), interstitial lung abnormalities (ILA), and idiopathic pulmonary fibrosis (IPF).^17–20^ Given the observed overlap in radiographic patterns and mechanisms of disease between COVID-19 ALI, ILA, and IPF, we hypothesized that biomarkers from the aforementioned classes would be associated with fibrotic pulmonary radiographic patterns in moderate to critical COVID-19 survivors during first three years following acute COVID-19 illness.

In a single-center New York City-based 3-year prospective longitudinal discovery cohort of adults hospitalized with severe and critical COVID-19 in 2020, we found that only higher levels of club cell secretory protein (CC16), encoded by the *SCGB1A1* gene, were consistently associated with an increased risk of fibrotic-like abnormalities in adjusted analyses. We then sought to externally validate the association of post-acute plasma CC16 levels and fibrotic radiographic patterns in two independent Canadian cohorts of adult moderate-to-critical COVID-19 survivors.

We tested the association between serum CC16 and larger airway-to-lung ratio on thoracic CT-scans at 15-month follow-up in our discovery cohort. To investigate the pulmonary source of elevated serum CC16 levels, we analyzed single cell RNA sequencing (scRNA-seq) and immunofluorescent staining of lung tissue from COVID-19 survivors.

## METHODS

### Study design and participants

The Columbia University Irving Medical Center (Columbia)-based discovery cohort has been described in detail previously.^7,13,21^ We prospectively enrolled ambulatory community-dwelling adults from New York City ages ≥21 years hospitalized at the Columbia tertiary-care Milstein Hospital and community-based Allen Hospital with laboratory-confirmed SARS-CoV-2 infection and severe or critical COVID-19 between March 1 and May 15, 2020. We excluded those with a history of interstitial lung disease or lung transplantation. We weighted sampling to include approximately 50% who required invasive mechanical ventilation during their COVID-19 hospitalization. We enrolled a total of 150 participants. Seventy-six participants were initially enrolled at 4-months after hospital discharge and invited to participate in 15-month and 3-year follow-up. We prospectively enrolled 47 and 27 additional participants with the same inclusion criteria and 50% sample weighting for requiring mechanical ventilation to increase and then maintain the sample size at ∼100 participants at 15-month and 3-year follow-up. In prior analyses, we found that demographic, genomic, comorbidity, and acute COVID hospitalization characteristics did not differ between those who did and did not follow-up across study visits, suggesting that cohort attrition was not affected by differential loss to follow-up.^13,21^

For all study cohorts, fibrotic-like abnormalities were defined as reticulations, traction bronchiectasis, or honeycombing.

We compared single cell analyses of transbronchial lung biopsies of fibrotic-like abnormalities in study participants obtained between 3 and 4 years after acute COVID-19 to transbronchial biopsies from four asymptomatic lung transplant recipients done as part of routine surveillance during the first year after lung transplantation. Since there was no evidence of infection or rejection and normal appearing lung architecture in the lung transplant biopsies, these samples were used as healthy lung tissue controls. We conducted immunofluorescence studies of explanted lungs from 7 adults who underwent lung transplantation for COVID-19 pulmonary fibrosis, and non-diseased lung tissue of 11 adults who underwent surgical resection of pulmonary nodules. A thoracic pathologist (AS) reviewed all transbronchial biopsies and explant lung tissues. All studies were approved by the Columbia Institutional Review Board (IRB) (AAAT5605, AAAT0009, and AAAS0753).

The University of British Columbia (UBC) validation cohort has been described in detail previously.^22^ Adults with COVID-19 hospitalized in Vancouver, Canada between March and May 2020 with laboratory-confirmed SARS-CoV-2 infection were prospectively enrolled with thoracic CT scans and plasma biobanking at 3 months. Patients with pre-existing interstitial lung disease were excluded. The study was approved by the UBC IRB (#H20-01239).

The McGill validation cohort was sampled as a nested case-control study cohort from the Biobanque Québécoise de la COVID-19 cohort (www.BQC19.ca), since only some participants received post-hospitalization thoracic scans for clinical care.^23^ We sampled adults hospitalized with laboratory confirmed SARS-CoV2 infection and COVID-19 from the Jewish General Hospital (JGH) in Montréal, Québec, Canada, excluding those with fibrotic-like abnormalities on thoracic CT scans within five years prior to the SARS-CoV-2 pandemic, interstitial lung disease, lung transplantation, and end-stage renal failure. Cases were defined as patients who were hospitalized with COVID-19 between March 2020 and November 2021 and had fibrotic-like abnormalities noted on clinical reports of thoracic CT scans assessed either within one week of hospital discharge, or approximately 4-months or 15-months after hospital discharge. Controls were sampled with the same inclusion criteria in a 2:1 ratio to cases and defined as patients with no mention of fibrotic-like abnormalities on CT scans obtained at similar follow-up intervals. After finding a higher prevalence of honeycombing than in the other cohorts, we re-reviewed the four cases with honeycombing on post-COVID thoracic CT. We conducted a post-hoc sensitivity analysis excluding three cases who were ultimately found to have possible honeycombing and lung distortion on thoracic CT done more than five years prior to the SARS-CoV-2 pandemic.

Columbia and UBC participants or a legally authorized representative signed written informed consent. Consent was obtained based on the BQC19’s standard operating procedures.^23^

### Thoracic Computed Tomography Analyses

For the Columbia discovery cohort, we obtained non-contrast high resolution CT scans at maximal inspiration at 4-months, 15-months, and 3-years after hospitalization for COVID-19. Two chest radiologists evaluated each thoracic CT as previously described.^7,13,24^ We examined 15-month thoracic CT scans to assess airway-to-lung ratio for patho-radiographic analyses. Airway lumen diameters were assessed at 19 standard anatomic locations (trachea to subsegments) and total lung volumes were segmented and measured from inspiratory thoracic CT images using Apollo Software (see E-Methods). We calculated airway-lung ratio using previously established methods.^25^

For the UBC validation cohort, we obtained high-resolution thoracic CTs. Two cardiothoracic radiologists independently evaluated radiographic abnormalities by separating each lung into three zones and estimating the percent of lung affected by either ground glass or reticulation. The presence versus absence of traction bronchiectasis and honeycombing was also noted in each lung zone. Further details have been described previously.^22^

In the McGill validation cohort, thoracic CT scans were obtained for clinical care. Fibrotic-like abnormalities were defined as a mention of reticulations, traction bronchiectasis, or honeycombing in the clinical report.

### Biobanking and Biomarker Analyses

For the Columbia and UBC cohorts, we prospectively obtained peripheral blood samples at follow-up. Hospital discharge samples for the Columbia cohort were obtained from remaining blood samples used for clinical care within 7 days of hospital discharge. The UBC cohort had no hospital discharge samples. For the McGill cohort, we selected prospectively collected plasma samples within 7 days of hospital discharge. The McGill cohort had no samples at post-hospitalization follow-up. Serum and plasma levels of biomarkers were assessed in the discovery and validation cohorts, respectively. Therefore, biomarker levels cannot be compared between discovery and validation cohorts. See E-Methods for further details.

### Single cell RNA sequencing analysis of Lung Transbronchial Biopsy Samples

We employed fluorescence-activated cell sorting (FACS) to separate and quantitate populations of lung cells prior to single cell RNA sequencing. Post-processing analysis was performed using Seurat v4.4.0.^26^ We employed a combination of an unsupervised and supervised approach to determine cell type annotation. We identified top genes that were unique to each cell cluster using *FindMarkers* using ROC analysis. See E-Methods for details.

### Immunofluorescence staining and image analysis of lung explant samples

Details of staining for SCGB1A1 and MUC5B and quality control are described in the E-Methods.

### Statistical Analyses

We examined unadjusted associations of biomarkers and clinical characteristics with Student’s T, Mann-Whitney, Chi-squared, Fisher exact, ANOVA, and Kruskal Wallis tests. We created separate generalized additive models (GAMs) with locally weighted smoothing to assess for nonlinear adjusted associations between biomarkers and the predicted risk of fibrotic-like abnormalities. We natural-log transformed right-skewed biomarker data. We used covariate balancing propensity scores (CBPS) to adjust for covariables to avoid overparameterized models at our study sample size, as we have done previously.^7,27^ Covariates included in CBPS were age, sex, race/ethnicity, body mass index, smoking history, COPD, asthma, estimated glomerular filtration rate, use of corticosteroids during COVID-19 hospitalization, IL-6 receptor inhibitor therapy during COVID-19 hospitalization, ventilator days during COVID-19 hospitalization, and days between SARS-CoV2 infection and thoracic CT scan. We estimated adjusted odds ratios per natural log-fold change in CC16 or by tertiles of CC16 using logistic regression. See E-Methods for further details.

## DATA AVAILABILITY

The data that support the findings of this study are available on request from the corresponding author (MRB). The data are not publicly available due to them containing personally identifiable information that could compromise research participant privacy and consent.

## RESULTS

### Participant characteristics, thoracic imaging, and biomarkers

There were 150 adults in the Columbia University discovery cohort with 76, 104, and 102 participants at 4-month, 15-month, and 3-year follow-up, respectively (**Figure 1A**). Forty-one (27%) participated in all three follow-up visits, 50 (33%) participated in two visits, and 59 (39%) participated in only one visit (**Figure E1**). Demographic and clinical characteristics did not appear to differ across follow-up visits (**Table E1**).^13^ There were 56 adults in the UBC validation cohort with 3-month follow-up, and 37 adults in the McGill validation cohort. The Columbia cohort was predominantly Black or Hispanic, and the UBC and McGill validation cohorts were mostly White or Asian. Compared to Columbia participants, UBC and McGill participants were slightly older, less obese, and had a lower prevalence of asthma. Compared to the UBC and McGill cohorts, the Columbia cohort had greater severity of acute COVID-19 illness, with more patients receiving mechanical ventilation (46% vs. 21% and 5.4%) and having a longer median [IQR] hospital length of stay (18 [7-42] vs. 8.5 [5.0-13] and 7 [5.0-19] days) (**Table 1**).

**Figure 1.**
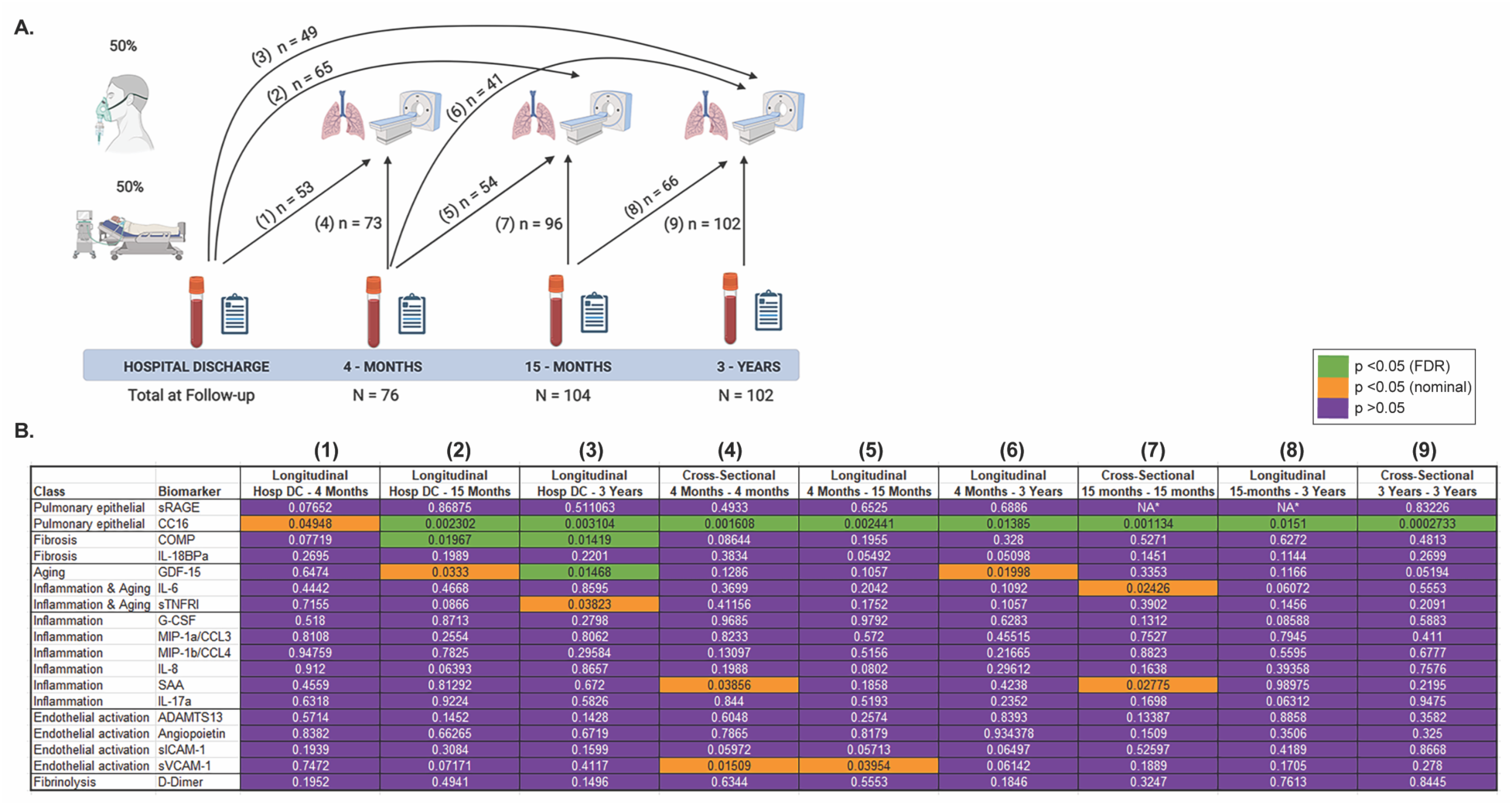
Discovery cohort study design and associations of biomarkers with fibrotic-like abnormalities. (**A**) Cohort study design of hospitalized survivors of severe COVID-19, with sampling weighted for 50% invasive mechanical ventilation survivors. Schema of measurements and associations of blood biomarkers and high-resolution chest CT scans with sample size available at each time point. (**B**) Associations of serum biomarkers with fibrotic-like abnormalities. P-values represent the p-for-association between the plasma biomarker and predictive risk of fibrotic pattern on thoracic CT using generalized additive models with LOESS smoothers, with adjustment for age, sex, race/ethnicity, BMI, COPD, pack-years history of smoking, estimated glomerular filtration rate, use of corticosteroids during COVID-19 hospitalization, IL-6 receptor inhibitor therapy during COVID-19 hospitalization, ventilator days, and days since initial SARS-CoV-2 infection utilizing covariate balanced propensity scores.

**Table 1.** Patient demographics, comorbidity, and acute COVID-19 illness severity measures.

| **Characteristics** | **Columbia**<br>**(Discovery)**<br>**cohort**<br>**(n = 150)** | **UBC**<br>**(Validation)**<br>**3-month follow-**<br>**up cohort**<br>**(n = 56)** | **McGill (Validation)**<br>**Hospital DC through**<br>**15-month follow-up**<br>**cohort**<br>**(n=37)** |
| --- | --- | --- | --- |
| Age, years, mean (SD) | 56 (13) | 61 (16) | 65 (15) |
| Male, n(%) | 83 (56%) | 33 (59%) | 15 (41%) |
| Body Mass Index (kg/m2),<br>mean (SD) | 33 (8.1) | 28 (16) | 29 (5.4) |
| Hispanic, n(%) | 100 (67%) | 1 (1.8%) | 3 (8.1%) |
| Race, n(%) |  |  |  |
| White | 55 (37%) | 25 (45%) | 27 (73%) |
| Black | 35 (23%) | 0 (0%) | 0 (0%) |
| Asian | 5 (3.3%) | 17 (30%) | 4 (11%) |
| Other | 54 (36%)<sup>a</sup> | 14 (25%)<sup>b</sup> | 6 (16%)<sup>c</sup> |
| Comorbidities, n(%) |  |  |  |
| Ever smoker | 58 (39%) | 18 (32%) | 10 (27%) |
| Active smoker | 4 (2.7%) | 2 (3.6%) | 1 (2.7%) |
| COPD | 7 (4.7%) | 2 (3.6%) | 1 (2.7%) |
| Asthma | 30 (20%) | 2 (3.6%) | 2 (5.4%) |
| Cardiovascular disease | 19 (13%) | 10 (18%) |  |
| eGFR, mean (SD) | 95 (45) | 81 (23) | 83 (26) |
| **Acute COVID-19 Illness Characteristics** |  |  |  |
| Admission SOFA score, median<br>[IQR] | 3 [2-5] | NA | NA |
| Corticosteroid use, n(%) | 67 (47%) | 2 (3.6%) | 23 (62%) |
| IL-6 receptor inhibitor use, n(%) | 31 (21%) | 2 (3.6%) | 1 (2.7%) |
| Oxygen use, n(%) |  |  |  |
| None | 0 (0%) | 14 (25%) | 7 (19%) |
| Nasal Cannula | 49 (32%) | 21 (38%) | 26 (70%) |
| Non-Rebreather | 28 (19%) | 0 (0%) | 0 (0%) |
| NIPPV or HFNO | 4 (2.7%) | 8 (14%) | 2 (5.4%) |
| Mechanical ventilation | 68 (46%) | 13 (21%) | 2 (5.4%) |
| Ventilator days, median [IQR] | 34 [14-46] | 8.5 [5.5-13] | 14 [12-24] |
| Hospital days, median [IQR] | 18 [7.0-42] | 8.5 [5.0-16] | 7.0 [5.0-19] |

UBC: University of British Columbia. DC: discharge. COPD: chronic obstructive pulmonary disease. eGFR: estimated glomerular filtration rate using the 2021 Chronic Kidney Disease (CKD)-EPI creatinine equation<sup>52</sup>. NIPPV: Non-invasive positive-pressure ventilation. HFNO: High-flow nasal oxygen. <sup>a</sup>4 Native American, 50 declined <sup>b</sup>13 South Asian, 1 mixed race. <sup>c</sup>4 Middle Eastern, 1 other, 1 unknown. For Columbia participants, body mass index is that recorded at first follow-up visit of the participant, eGFR is recorded from hospital discharge, and active smoker is active smoking reported at any follow-up visit.

The prevalence of fibrotic-like abnormalities at follow-up in the Columbia and UBC cohorts ranged from 57-64%, and was 43% in the nested case-control cohort from McGill, with 16 participants with and 21 participants without fibrotic-like abnormalities, matching on demographics, comorbidities, and either hospital discharge, 4-month, or 15-month thoracic CT follow-up time. Fibrotic-like abnormalities were predominantly characterized by reticulations and traction bronchiectasis. We observed honeycombing in <5% of Columbia and UBC patients, and in 5 (14%) McGill patients (**Tables E2 and E3**).

In the Columbia cohort, of the 18 biomarkers tested, only club-cell secretory protein-16 (CC16) had significant associations with the predicted risk of fibrotic-like abnormalities in adjusted analyses (**Figure 1B**). Sensitivity analyses revealed potential model overfitting with high outliers after natural-log transformation observed in six end-stage renal disease (ESRD) dialysis patients. Since CC16 is renally excreted,^28,29^ we report CC16 analyses excluding Columbia participants with ESRD and excluded ESRD patients in validation cohorts. CC16 declined slightly over three years but remained consistently higher among those with fibrotic-like abnormalities (**Figure 2A**). In nine cross-sectional and longitudinal adjusted associations tested between hospital discharge and 3-year follow-up, CC16 had consistent, large magnitude, direct, and mostly linear associations with the predicted risk of fibrotic-like abnormalities (**Figure 2B**). The adjusted odds of fibrotic-like abnormalities for every natural log-fold increase in CC16 ranged from 5.0 (95%CI: 1.6-20.7) to 16 (95%CI: 4.1-111). Adjusted odds ratios increased two to seven-fold across tertiles of CC16 (all p-for-trend <0.02) (**Table E4**). Higher CC16 had a moderate inverse correlation with lower percent-predicted diffusion capacity of carbon monoxide (DLCO) at 4 months and 3 years but had no statistically significant correlation with DLCO at 15 months (**Table E5**).

**Figure 2.**
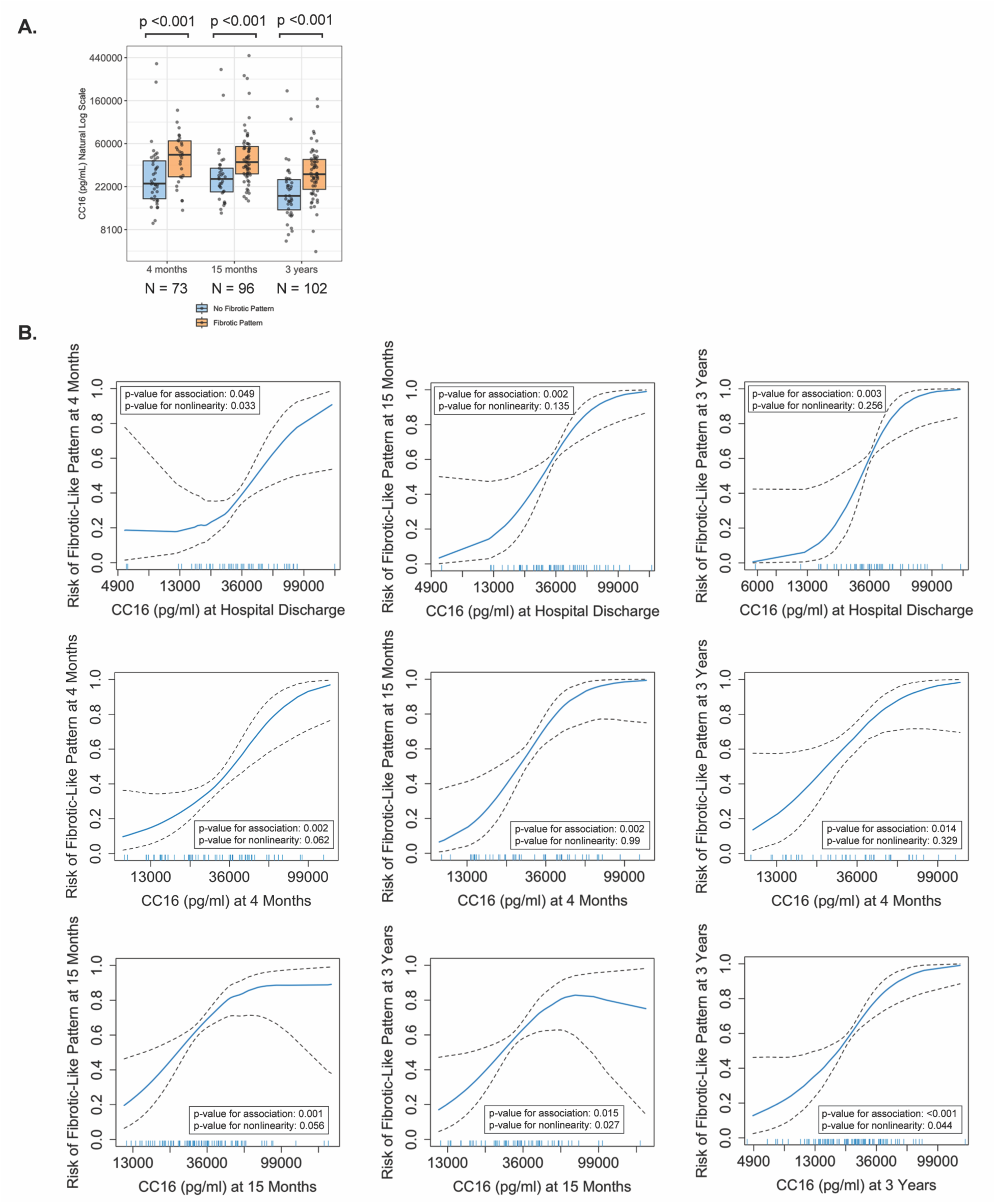
Associations of CC16 with fibrotic-like abnormalities in the discovery cohort. (**A**) Dot-box plots of plasma CC16 at hospital discharge, 4-months, 15-months, and 3-years, stratified by fibrotic pattern status in the Columbia (discovery) cohort. Boxes represent the interquartile range, and the middle bar represents the median. (**B**) Plots of generalized additive models (GAM) with LOESS smoothers with adjustment for age, sex, race/ethnicity, BMI, COPD, asthma, pack-year history of smoking, estimated glomerular filtration rate, use of corticosteroids during COVID-19 hospitalization, IL-6 receptor inhibitor therapy during COVID-19 hospitalization, ventilator days, and days since initial SARS-CoV-2 infection using covariate balanced propensity scores.

In the UBC cohort, 3-month cross-sectional analyses revealed a direct, linear, borderline significant association between CC16 and the predicted risk of fibrotic-like abnormalities (GAM p=0.058) that was statistically significant in unadjusted (p=0.037) and adjusted logistic regression analyses (OR, 95% CI per natural log fold increase in CC16: 2.92, 1.07-9.07) (**Figure 3, Table E6**). Higher CC16 had a moderate inverse correlation with lower percent-predicted DLCO (**Table E5**).

**Figure 3.**
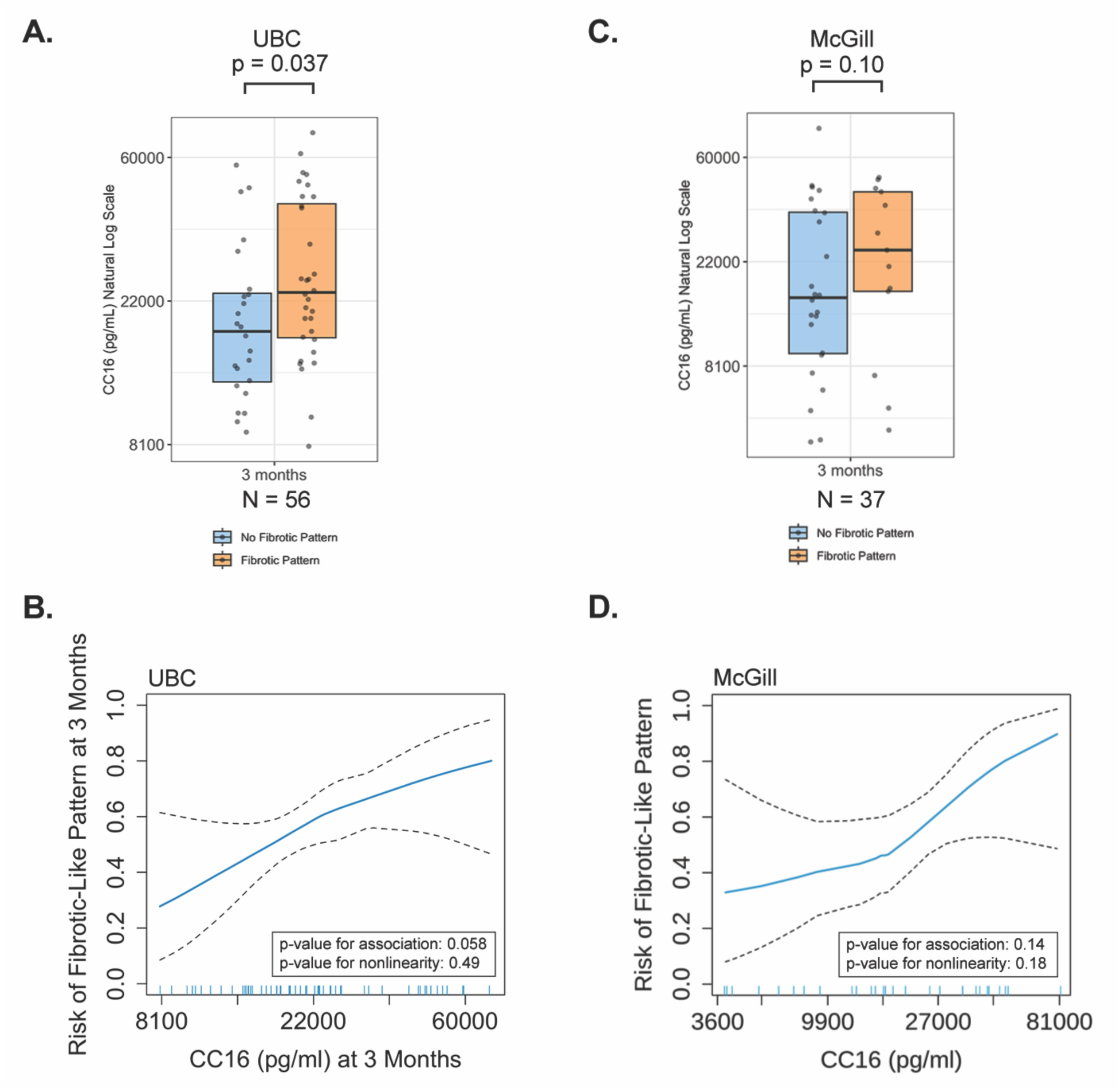
Associations of CC16 with fibrotic-like abnormalities in the validation cohorts. (**A, C**) Dox-box plots of plasma CC16 stratified by fibrotic pattern status in the University of British Columbia (UBC) and McGill University validation cohorts. Boxes represent the interquartile range and the middle bar represents the median. (**B, D**) Plots of generalized additive models (GAM) with LOESS smoothers with adjustment for age, sex, race/ethnicity, BMI, COPD, asthma, pack-year history of smoking, estimated glomerular filtration rate, use of corticosteroids during COVID-19 hospitalization, IL-6 receptor inhibitor therapy during COVID-19 hospitalization, ventilator days, and days since initial SARS-CoV-2 infection using covariate balanced propensity scores.

Given the small sample size of the McGill cohort, we tested associations and estimated effect sizes of CC16 with fibrotic-like abnormalities for the hospital discharge, 4-month, and 15-month follow-up participants combined. We observed a similar direct, linear, but non-significant association in unadjusted and adjusted analyses (**Figure 3**), with an adjusted odds of fibrotic-like abnormalities of 2.36, 95% CI: 0.93-7.03 per natural log-fold increase in CC16 (**Table E6**). In unadjusted analyses stratified by follow-up time period, we observed nonsignificant higher median CC16 levels in those with fibrotic-like abnormalities (**Figure E2**). In post-hoc sensitivity analyses excluding three patients with possible honeycombing found on thoracic CT greater than five years prior to the 2020 SARS CoV2 pandemic, we observed the magnitude of the associations to be slightly less, with the data still collectively suggesting an association of higher CC16 levels with fibrotic-like abnormalities, even though the sample size becomes small in analyses stratified by follow-up time period (**Table E6, Figure E3**).

In radiographic analyses of the Columbia cohort, higher levels of CC16 were directly and linearly associated with a higher airway-to-lung ratio at 15-months in adjusted longitudinal and cross-sectional analyses (**Figure 4**), with consistent effect sizes over time. For example, every natural log-fold increase in CC16 at hospital discharge was associated with a 0.61 (0.21-1.01) standard-deviation unit increase airway-to-lung ratio (**Table E7**). Associations were robust to complete-case and larger airway (pre-subsegmental) sensitivity analyses (**Figure E4**).

**Figure 4.**
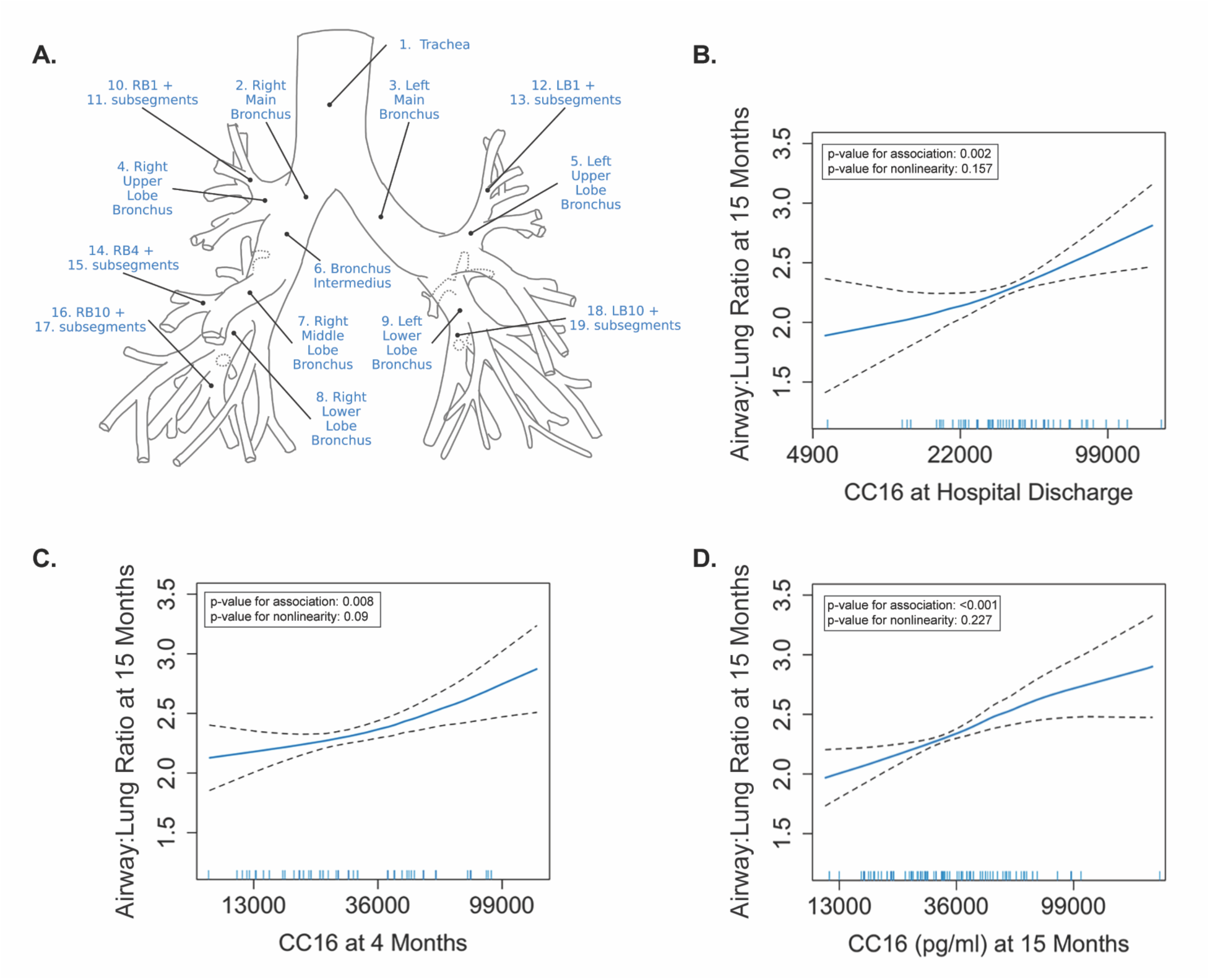
Associations of CC16 with airway-to-lung ratio measured on 15-month thoracic CT in the discovery cohort. (**A**) Using Apollo Software (VIDA diagnostics, Coralville, Iowa), airway to lung ratio was quantified on inspiratory thoracic CT performed at 15-months as the geometric mean of airway lumen diameters measured at 19 standard anatomic locations divided by the cube root of lung volume. (**B-D**) Plots are of generalized additive models (GAM) with LOESS smoothers with adjustment for age, sex, race/ethnicity, BMI, COPD, asthma, pack-year history of smoking, estimated glomerular filtration rate, use of corticosteroids during COVID-19 hospitalization, IL-6 receptor inhibitor therapy during COVID-19 hospitalization, ventilator days, and days since initial SARS-CoV-2 infection using covariate balanced propensity scores.

### Functional analyses with lung tissue

The demographics and clinical characteristics of four COVID-19 survivors with the highest fibrosis scores on thoracic imaging at 3-year follow-up^24^ and four control subjects (lung transplant recipients without histopathologic evidence of rejection or infection, **Figure E5**) are described in **Table E8**. A single cell suspension of each transbronchial lung biopsy sample was fluorescence-activated cell sorted to mix epithelial, immune/endothelial, and non-epithelial/non-immune/non-endothelial cells in a 1:1:1 ratio prior to sequencing. Clustering and distinction of epithelial cell types by the expression of top genes shows two groups of cells expressing *SCGB1A1* (**Figures 5B-5D**). The first, designated as SCGB1A1+MUC5B+ airway cells, do not express surfactant genes but robustly express mucin genes; these are secretory cells (**Figure 5E**). The second, designated as SCGB1A1+SCGB3A2+SFTPB+ airway cells, are similar to the previously described pre-terminal bronchiole secretory cells (pre-TB-SC) or respiratory airways secretory (RAS) cells (**Figure 5E**, **Figure E6**).^30,31^ There is evidence of increased proportions of both these SCGB1A1 expressing cell clusters when analyzed separately (**Figure 5F**) or in combination (**Figure 5G**) in COVID-19 samples as compared to control samples. Proportions of SCGB1A1 expressing cell clusters were more dissimilar between COVID-19 and control samples than the proportions of alveolar type 1 and type 2 epithelial cells (**Figure 5H**), suggesting that post-COVID effects drive expansion of airway rather than alveolar cells. Differences in fibroblast, immune, and endothelial populations between COVID-19 and control samples are shown in **Figure E6**. None of the COVID-19 cases carried the MUC5B risk allele that has been linked to pulmonary fibrosis.

**Figure 5.**
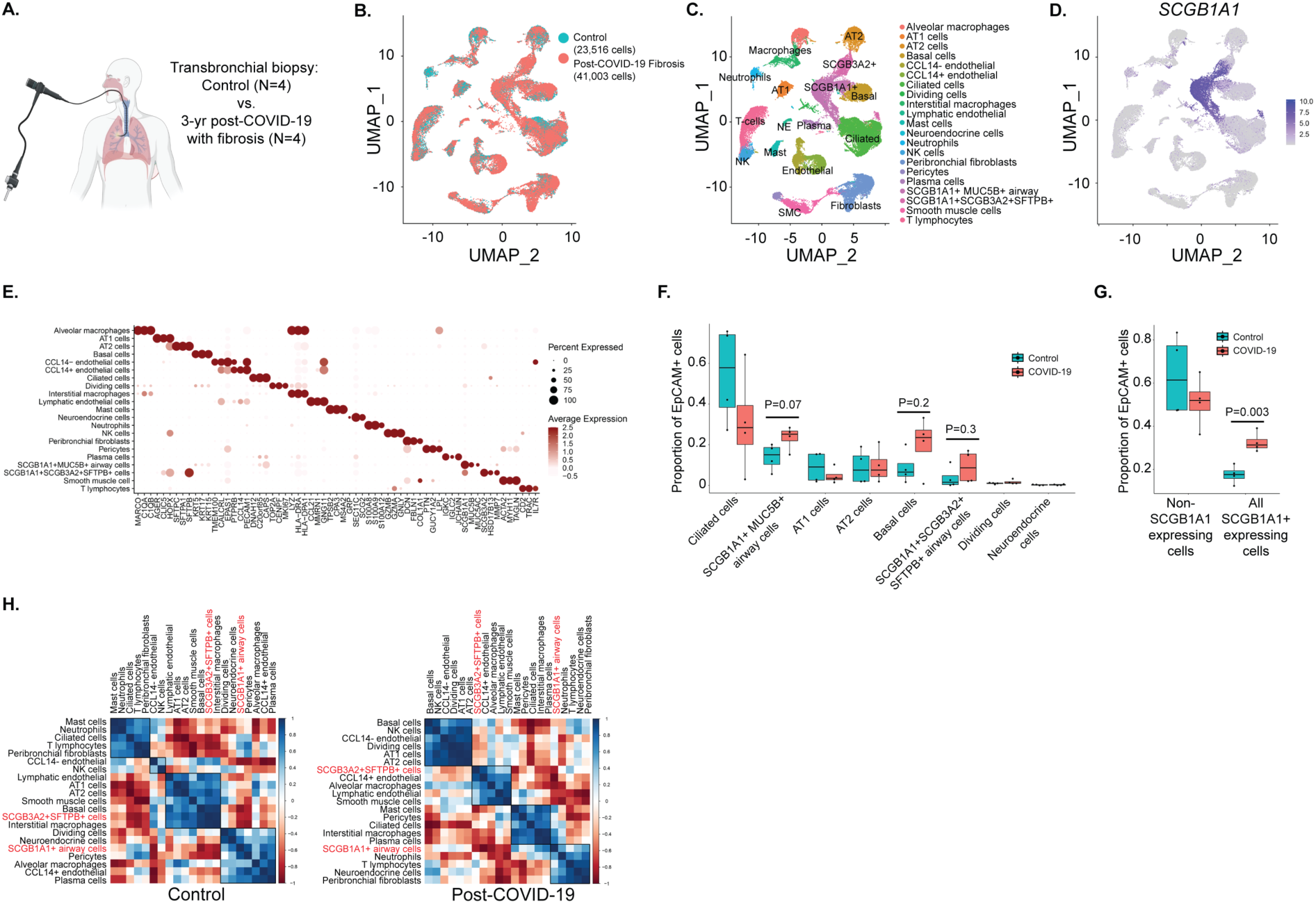
Increased proportions of SCGB1A1-expressing lung cells after severe COVID-19. (**A**) Schematic. (**B**) UMAP plot of cells from subjects with control lung (aqua) and post-COVID with fibrosis (red). (**C)** UMAP plot of annotated cell types. (**D)** Feature plot of *SCGB1A1* expression. (**E**) Dot blot plot of top expression genes in each of the different cell clusters. (**F**) Proportion of different subsets of lung epithelial (EpCAM+) cells found in control and COVID-19 with fibrosis. (**G**) Proportion of SCGB1A1+ expressing cells (from the SCGB1A1+MUC5B+ airway and SCGB1A1+SCGB3A2+SFTPB+ airway cell clusters) and non-SCGB1A1 expressing cells (all other EpCAM+ cell clusters) found in controls and COVID-19 with fibrosis. (**H**) Proportions of SCGB1A1+ expressing cells and other cell types in controls and COVID-19 with fibrosis samples.

We compared lung explants from seven individuals who underwent lung transplantation 2-29 months after COVID-19 (**Table E9**) to eleven normal control samples (lung resected adjacent to a pulmonary nodule). There was no difference between immunofluorescence SCGB1A1/CC16 staining of epithelial cells lining large (>100 micron) airways, but the post-COVID lung explants showed dramatically increased expression in low cuboidal epithelium lining small (<100 micron) airways (**Figure 6C, D**). Compared to control samples, COVID-19 explant samples (**Figure 6E**) had over a 3-fold increase in CC16-MUC5B co-expression (0-14% vs 18-26% cells, respectively, [p =0.0006]) (**Figure 6F, G**). These double positive cells were typically located in small airways in areas of peribronchial metaplasia, with MUC5B staining the cell apices and extracellular spaces, and CC16 staining the basolateral portion of the cells. Photomicrographs of immunofluorescence antibody controls are shown in **Figure E7**.

**Figure 6.**
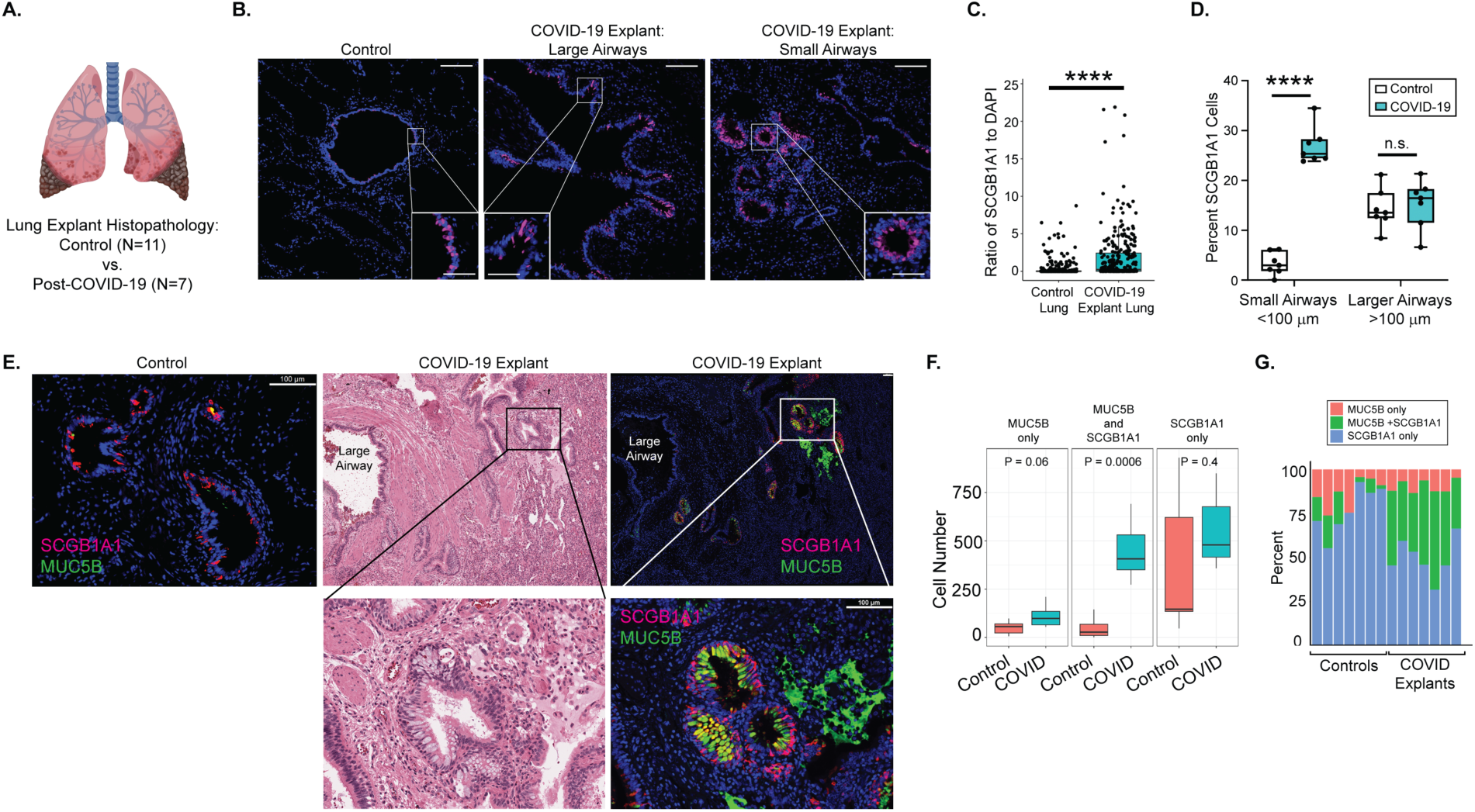
Increased SCGB1A1 immunofluorescence with MUC5B in small airways of the lung after severe COVID-19. (**A**) Schematic. (**B**) Immunofluorescence pattern of SCGB1A1 in large and small airways in control and COVID-19 explants (bars are 200 microns). Insets show staining at higher magnification (bars are 50 microns). (**C**) Automated quantification of SCGB1A1 immunofluorescence in control (n=11) and COVID-19 explanted lungs (n=7). Each point represents the average SCGB1A1+ areas divided by number of DAPI-stained cells of 42 photomicrographs for each case at 20X magnification (p < 2.2^e-16^). (**D**) Manual counting of SCGB1A1+ airway epithelial cells in airways measuring less than or greater than 100 µm in diameter in four slides from all n=7 COVID-19 explants and n=7 sex-matched controls. Small (<100 micron) airways p <1^e-6^; large airways p=0.67. (**E**) Representative co-immunofluorescent staining of SCGB1A1 and MUC5B in control and COVID-19 explant tissue (bars are 100 microns). Hematoxylin and eosin-staining of an adjacent cut from the COVID-19 explant is shown for reference. Higher power magnification images of the hematoxylin and eosin-stained slide and the immunofluorescent image from the areas denoted by the boxes are shown below. . MUC5B signals (green) are seen not only in cells but also within the adjacent extracellular space. (**F**) Quantification of the absolute number of cells staining with MUC5B alone, SCGB1A1 alone, or both MUC5B and SCGB1A1 in COVID-19 explants (n=7) and control lung tissue (n=7). (**G**) Bar graph demonstrating the percentage of airway cells with the indicated staining pattern found in COVID-19 explants (n=7) and control lung tissue (n=7).

## DISCUSSION

In three racial and ethnically diverse cohorts of adults hospitalized during the wild type and delta waves of COVID-19 pandemic, we found consistent cross-sectional and longitudinal associations of higher circulating CC16 associated with fibrotic-like abnormalities on thoracic CT for up to three years after hospitalization. CC16 appears to have a dose-response association with larger airway-to-lung ratio suggesting that its secretion may be proportional to cellular changes from SARS-CoV-2 mediated airway injury and repair. Our transcriptomic and immunofluorescent analyses confirm increased numbers of small airway epithelial cells expressing SCGB1A1/CC16 and co-expressing MUC5B in patients with post-COVID-19 fibrotic lung abnormalities. Collectively, our findings suggest that CC16 is a post-acute circulating biomarker of COVID-19 related fibrotic pulmonary patterns that represents pathologic remodeling of small airway epithelium leading to a pro-fibrotic state.

Human proteome maps show that CC16 expression occurs primarily in the lung epithelium. Experimental and clinical studies demonstrate that CC16 is a pleotropic negative regulator of inflammation. While the mechanisms by which CC16 decreases lung inflammation are not completely understood, CC16 attenuates acute inflammation via limiting neutrophil chemotaxis and limits chronic inflammation via inhibition of osteopontin, which stimulates epithelial cell production of T helper-cell 2 cytokines.^32,33^ Consistent with these experimental studies, higher levels of circulating CC16 are associated with less severe COPD and a slower decline in FEV1.^34^ Increased CC16 is observed in non-COVID ARDS and in severe COVID-19,^35,36^ presumably reflecting lung epithelial cell injury and alveolar-blood barrier leakage. Higher circulating CC16 levels are also found in multiple restrictive interstitial lung diseases,^32^ including idiopathic pulmonary fibrosis (IPF),^37,38^ where it may reflect a response to ongoing lung injury and repair. Additionally, our observation of an inverse correlation between CC16 and DLCO suggests that the increase in SCGB1A1 cells is linked to functional limitations, including decreased diffusion at the alveolar capillary interface. Our investigation is consistent with these prior studies insofar as CC16 levels are highest at hospital discharge, closest in time to COVID-19 acute lung injury. Its gradual decline but persistent relative elevation in those with fibrotic-like abnormalities on thoracic CT is consistent with the findings of restrictive interstitial lung disease studies.

The two SCGB1A1+ epithelial cell clusters identified by unsupervised clustering have been characterized as airway progenitor cells.^39,40^ The SCBG1A1+SCGB3A2+SFTPB+ cell cluster, termed a respiratory airway secretory (RAS) cell, can self-renew and generate alveolar type 2 cells.^30,31^ Similarly, SCGB1A1+ secretory cells can self-renew and contribute to airway epithelial repair after injury.^41^ The subset of SCGB1A1 cells that co-expresses MUC5B are normally found by immunohistochemistry to occupy the mid-portion of the human airway tree.^30^ But in this study, we find ∼3-fold more SCGB1A1+MUC5B+ cells anomalously located in the small (<100 micron) respiratory bronchioles in post-COVID-19 fibrosis. Interestingly, a recent scRNA-seq analysis also revealed enriched populations of secretory cell clusters with transcriptional signatures that include *SCGB1A1*, *SCGB3A2*, and *MUC5B* in IPF,^42^ suggesting that dual activation of alveolar and secretory epithelial repair programs are characteristic not only of IPF but also post-COVID fibrosis.

The rs3570590 MUC5B promoter variant confers an inherited risk factor for IPF and other types of pulmonary fibrosis by increasing MUC5B expression.^43–47^ This study finds large numbers of SCGB1A1+MUC5B+ secretory epithelial cells that persist years after the initial SARS-CoV-2 infection and aberrantly reside in elongated respiratory bronchioles, thus activating a pro-fibrotic program in the most distal aspects of the human airway tree. Although the COVID-19 survivors with fibrosis did not carry this genomic risk allele, we find evidence for increased numbers of SCGB1A1+MUC5B+ cells by immunofluorescence suggesting that viral infection may lead to acquired MUC5B overexpression. Thus, there may be convergence of genetic and virally-induced MUC5B overexpression that both lead to an increased risk of fibrosis. As this variant is associated with decreased hospitalization and improved survival after COVID-19,^2,48^ it may have been subject to natural selective pressure from preceding waves of deadly respiratory viral infections leading to its enrichment in European populations.

The identification and scoring of fibrotic-like residual abnormalities by thoracic radiologists, particularly traction bronchiectasis, can be difficult and subjective.

Therefore, we took the novel approach of validating a dose-response association of CC16 with fibrotic-like abnormalities using airway-to-lung ratio calculated from supervised machine-learning software. Previously, lower airway to lung ratio, as an index of size dysanaptic lung growth revealed an association with COPD risk among community-dwelling older adults.^25^ Our finding that higher circulating CC16 levels are linearly associated with greater airway-to-lung ratio at 15-months suggests that the level of circulating CC16 may reflect extent post-infectious remodeling of airway architecture as evidenced by shifts in the proportions of epithelial cells observed in our transcriptomic analyses, and the robust expression of SCGB1A1 in the small airways of COVID-19 survivors in our histopathologic analyses. These lung architectural changes may manifest as traction bronchiectasis or bronchiolectasis, and an increased airway-to-lung ratio.

Our study has limitations. The sample sizes of our study cohorts are modest, ranging in size from 37 to 150 participants. Accordingly, we lack statistical power to detect statistically significant association in the smallest validation cohort. Each cohort had differences in demographics, the prevalence of invasive mechanical ventilation use, and methods to assess for persistent lung abnormalities on chest CT, which likely account for some variation in the observed prevalence of reticulations and traction bronchiectasis across cohorts. Despite these differences, our consistent associations of CC16 with fibrotic-like abnormalities suggest that our results are robust to variation in case-mix and measurement. The observed effect sizes in the discovery cohort may overestimate what would be observed in a larger population of COVID-19 survivors since we oversampled invasive mechanical ventilation survivors, who likely had the highest CC16 levels acutely and who are at the greatest risk for post-COVID residual fibrotic lung abnormalities.^7^ These findings underscore the importance of following COVID-19 ARDS survivors in the future. We did not test associations of pulmonary epithelial cell proteins, such as metalloproteinase-7 (MMP-7), Krebs von den Lungen-6 (KL-6), surfactant protein-A (SPA), and surfactant protein-D (SP-D) that have been linked to IPF, and some of which have recently been found to be associated with reduced lung diffusion capacity in COVID-19 survivors.^49^ We could not confirm whether the “control” transplanted lung or the lung resections had prior SARS CoV-2 infection that resolved without radiographic or histopathologic abnormalities. While the inclusion of lung transplant recipients as controls for the single cell analysis may have had transcriptional alterations related to their immunosuppression medications, the findings relevant to CC16/SCGB1A1 were also replicated in the immunofluorescent analysis of explant tissue and control lung resections adjacent to nodules in non-immunosuppressed individuals. Thus, the persistent transcriptomic and immunohistochemical changes that we identified in COVID-19 survivors with fibrotic-like abnormalities may reflect other sequelae of SARS-CoV-2 infection itself. The numbers of subjects and tissue available for the single cell RNA and immunohistochemistry analyses, although age- and sex-matched, are limited and may be subject to sampling bias. The prevalence and extent of fibrotic changes one or more years after non-COVID ARDS in the low tidal volume ventilation era is less that observed in COVID-19 ALI/ARDS,^50,51^ but is still occasionally observed. Whether our finding of elevated CC16 and dysregulated airway epithelial progenitor cell remodeling is specific to COVID-19 or a generic response in all ALI/ARDS survivors with fibrotic changes remains unknown.

Cohort enrollment occurred primarily during the wild-type wave of the SARS-CoV2 pandemic, with some McGill participants sampled from the early Delta strain wave. Whether our findings remain consistent with vaccination, subsequent SARS-CoV2 variant infection remains unknown. Future studies in larger population-based cohorts are needed to investigate the potential clinical utility of using CC16 levels to prognostically enrich and facilitate screening of COVID-19 survivors for fibrotic lung abnormalities.

In conclusion, CC16 remains consistently elevated and associated with thoracic CT fibrotic-like abnormalities for 3-years post-hospitalization among adult survivors of moderate to critical COVID-19. Elevated circulating CC16 appears to reflect underlying deranged pulmonary epithelial progenitor proliferation and anomalous CC16/MUC5B-related fibrotic signaling in distal human airways. These findings suggest that CC16 should be investigated further as a blood biomarker that may be leveraged to facilitate screening of COVID-19 survivors for residual fibrotic lung abnormalities and their functional consequences.

## Data Availability

All data produced in the present study are available upon reasonable request to the authors, and after joint institutional IRB approval of a data sharing agreement.

## ACKNOWLEDGEMENTS

We are grateful to all participants and their families. This study would not be possible without their time and support of research. We are grateful for the technical expertise of Lesley Vickers, Dr. Niyati Desai, and Dr. Michael Miller, Director of Digital Pathology. BioRender was used to create schematic diagrams.

## AUTHOR CONTRIBUTIONS

The manuscript was initially drafted by MRB, with support from CKG. MRB, CKG, CC, CJR, JBR, and TN made substantial contributions to the conception and design of the work. AEJ, SOM, CFM, CKG, ACYY, PJ, AWW, ASS, and NK have accessed and verified data of the respective cohorts. RN had oversight of the plasma and serum biomarker assessments. All authors contributed to data interpretation, critical review and revision of the manuscript, and final approval of the version to be published. All authors are responsible for the decision to submit the manuscript and are accountable for all aspects of the work in ensuring that questions related to the accuracy or integrity of any part of the work are appropriately investigated and resolved.

## MATERIALS AND CORRESPONDENCE

Data sharing will require an IRB-approved data sharing agreement. Contact Matthew R. Baldwin, MD, MS at for Columbia University discovery cohort data inquires, Christopher Carlsten, MD, MPH at for University of British Columbia validation cohort data inquires, and Tomoko Nakanishi, MD, PhD at for McGill data from Biobanque Québécoise de la COVID-19 cohort data inquires.

