## Supplementary material for "Pulmonary fibrosis after COVID-19 is characterized by airway abnormalities and elevated club cell secretory protein-16": E-Tables and E-Figures

**TABLE OF CONTENTS (in order of citation in the main manuscript)**

**E-Methods**

**Figure E1.** Hematoxylin and eosin staining of transbronchial biopsies representing healthy lung tissue for single-cell analyses that were taken from lung transplant recipients in the first year after lung transplantation for routine surveillance rather than symptomatic evaluations.

**Figure E2**. Columbia (discovery) cohort follow-up visit participation.

**Table E1**. Demographic and clinical characteristics stratified by follow-up visit in the Columbia (discovery) cohort.

**Table E2**. Fibrotic-like abnormalities on Thoracic CT scans in the Columbia (discovery), University of British Columbia (validation), and McGill (validation) cohorts.

**Table E3**. Comparison of demographic and clinical characteristics between those with and without fibrotic-like abnormalities in the McGill (validation) nested case-control cohort.

**Table E4**. Effect estimates of associations between CC16 and fibrotic-like lung abnormalities on thoracic CT scan in the Columbia (discovery) cohort.

**Table E5**. Cross-sectional spearman correlations of CC16 and percent predicted diffusion capacity of lung for carbon monoxide (DLCO) at follow-up in the Columbia (discovery) and University of British Columbia (validation) cohorts.

**Table E6**. Effect estimates of associations between CC16 and fibrotic-like lung abnormalities on thoracic CT scan in the University of British Columbia and McGill (validation) cohorts.

**Figure E3**. Circulating CC16 (pg/ml) levels in those with and without fibrotic-like abnormalities in the McGill (validation) nested case-control cohort, stratified by follow-up time period of thoracic CT scan: (A) hospital discharge, (B) 4 months; (C) 15 months.

**Table E7.** Generalized linear model adjusted associations of CC16 with airway-to-lung ratio 15 months in Columbia (discovery) cohort.

**Figure E4.** Circulating CC16 (pg/ml) levels in those with and without fibrotic-like abnormalities in the McGill (validation) nested case-control cohort sensitivity analysis excluding n=3 case patients with honeycombing in the McGill cohort (A), and stratified by follow-up time period of thoracic CT scan: (B) hospital discharge, (C) 4 months; (D) 15 months. Sensitivity analysis. Generalized additive model (GAM) with LOESS smoothers with adjustment for age, sex, race/ethnicity, BMI, COPD, pack-year history of smoking, estimated glomerular filtration rate, ventilator days, and days since initial SARS-CoV-2 infection using covariate balanced propensity scores.

**Figure E5**. Sensitivity analyses of generalized additive model adjusted associations between CC16 and airway to lung ratio. (A-C) Complete case analyses among those with all subsegmental airways visualized. (D-F) Analyses of all subjects excluded sub-segmental measurements.

**Table E8.** Demographics and clinical characteristics of COVID-19 survivors with fibrosis on thoracic imaging at 3-year follow-up and lung transplant receipts who underwent transbronchial biopsies

**Figure E6**. Single cell RNA sequencing quality control, gene expression data, and differences in cell numbers of immune, endothelial, and non-epithelial, non-endothelial, and non-immune cells

**Table E9.** Demographics of patients who underwent lung transplantation for COVID-19 pulmonary fibrosis and controls who underwent surgical resection of pulmonary nodules.

**E-References**

### E-METHODS

#### Thoracic CT Airway-to-Lung Ratio analyses

Apollo software trained technologists were unaware of other participant information. At least one subsegmental lumen (see Figure 4) either did not exist or was not visualized by the Apollo software among 22 of 65 15-month participants with CC16 measured at hospital discharge, 14 of 54 15-month participants with CC16 measured at 4 months, and 32 of 96 15-month participants with CC16 measured at 15 months. In our primary analysis (Figure 4), we report the geometric mean allowing for missing subsegmental values. We conducted two sensitivity analyses to ensure that the observed results are robust to small airway detection bias. (1) We conducted a complete-case analysis not allowing for missing subsegmental values. (2) We assessed the geometric mean from trachea to segments, excluding subsegments entirely.

#### Biobanking and Biomarker Analyses

For all cohorts, prospectively collected blood samples were centrifuged immediately, and supernatant was aliquoted and cryopreserved at -80°C.

Among Columbia participants who co-enrolled in the Columbia University COVID-19 Biobank initiative (IRB AAAS7370), we used the sample closest in time to the date of hospital discharge with a limit of +/- 10 days of discharge (observed median[IQR] -2 [-3.5 to -1] days). Among McGill participants, we assessed the plasma sample closest in time to the date of hospital discharge, with a limit of +/- 10 days (observed median[IQR] -2 [-1 to -7]).

All biomarker assessments were performed in duplicate at the Columbia University Irving Institute for Clinical and Translational Research using Miliplex multiplex assays with the Luminex xMAP bead-based multiplex assay platform (Milipore Sigma, USA). We assessed biomarkers from the soluble cytokine receptor panel, human cardiovascular panel 2, human cytokine/chemokine/growth factor Panel A, human angiogenesis/growth factor 1, and human fibrosis panel (which includes club cell secretory protein-16 [CC16], also known as uteroglobin). For CC16, the median [IQR] coefficients of variation for CC16 were 3.3% [1.8-6.6%], 4.5% [2.4-9.7%], 3.3% [1.8-6.6%], and 4.6 [2.2-10%]) at hospital discharge, 4-month, 15-month, and 3-year Columbia cohort samples, and 5.2% [2.4-9.2%] and 5.4% [3.1-8.4%] for the UBC and McGill cohort samples, respectively.

#### Fluorescence-activated cell sorting (FACS) of Lung Transbronchial Biopsy Samples

A cell suspension was prepared from transbronchial lung biopsies by digestion in 1.4ml DMEM containing 0.3 mg/µl liberase (Sigma), 70 µg/µl elastase (Worthington) and 3 U/ml dispase II (Sigma) at 37°C for 45 min, followed by the addition of 10µl DNAse (2.7 U/µl; Qiagen), 1µl 1M DTT, and 2µl 0.5M EDTA, and incubation for an additional 15 min. The cell suspension was strained through a 100-µm filter, centrifuged at 300*xg* for 10min, and resuspended in ~200µl residual buffer plus 10µl DNAse. Red blood cells were lyzed by adding 1ml ACK buffer (Gibco) and incubating on ice for 2 min. Following the addition of 10ml PBS with 1% BSA, centrifugation and resuspension in ~200µl residual buffer plus 10µl DNAse, the cells were blocked with FcR Blocking Reagent (Miltenyi) and stained 1:100 with CD326(EpCAM)-AF488 (Invitrogen), CD31-APC (Biolegend), CD45-APC (Biolegend), and CD235a-PE (Biolegend) for 1hr at 4 $^{o}$C, strained through a 100-µm filter, and flow sorted on a BD Influx fluorescence-activated cell sorter (FACS). We positively sorted for complex, singlet cells and negatively sorted for DAPI and PE, which removed the dead cells and any remaining red blood cells. The FACS-sorted cells were collected as three populations: CD326/EpCAM positive (epithelial), CD31/CD45 positive (immune and endothelial), and those negative for either AF488 or APC fluorophore signals (enriched for fibroblasts). Using the cell count numbers obtained by FACS, each of the three populations of cells were mixed 1:1:1, using the population with the fewest number of cells as the limiting factor. Following centrifugation and resuspension in PBS with 0.04% BSA, cell viability was measured using trypan dye exclusion, and 10,000 cells were loaded on a 10X Genomics flow cell for encapsulation in individual droplets for lysis, reverse transcription, and barcoding of cDNA, followed by sequencing on an Illumina NovaSeq 6000. We performed additional quality control by visualizing transcript count, gene count, and percent mitochondrial genes for each cell cluster and by each sample.

**Genotyping**

DNA was isolated from each Columbia participant at follow-up and DNA was isolated from blood leukocytes using the Gentra Puregene Blood Kit (Qiagen, Valencia CA). Geotyping for the *MUC5B* rs35705950 risk allele was obtained by Sanger sequencing.

**Single cell RNA sequencing analysis**

Cells were filtered using *Subset* function to remove those with >10% mitochondrial genes and <200 or >7500 genes expressed and <4000 or >40000 transcripts identified. Sample-level datasets were integrated using reciprocal PCA. *NormalizeData* function was used to log normalize data and the *ScaleData* function was used to regress out the number of UMIs and percentage of mitochondrial genes. Principal component analysis was performed using *RunPCA* and Louvain clustering was performed using the *FindNeighbors* and *FindClusters* functions. UMAP embeddings were calculated using *RunUMAP* for visualization. Correlation plots of the proportion of cell clusters in control and COVID-19 samples were graphed using R package *CorrPlot.*

To interpret cellular compositions of each sample, we grouped all annotated cell types by FACS groups corresponding to epithelial cells (EpCAM+), endothelial/immune cells (CD31+/CD45+) and stromal or other cells (EpCAM-/CD31-/CD45-). We visualized the transcriptomic expression of *EPCAM*, *PTPRC* (CD45) and *PECAM1* (CD31) in each cell type to confirm correct membership.

**Immunofluorescence Staining**

Formalin-fixed paraffin-embedded (FFPE) lung slides were dewaxed using xylene (three times, 5 minutes each), rehydrated using gradients of ethanol (100, 95, 70, and 50% ethanol; two times, 10 minutes each), and washed with deionized water (two times, 5 minutes each). For quantification of SCGB1A1 immunofluorescence, antigen retrieval was performed by immersing the slide into 10mM Sodium Citrate buffer (pH 6.0) at 95˚C and then cooling to room temperature for 30 minutes. The slides were washed in distilled water for 5 minutes, permeabilized by 0.1% Triton-X 100 in TBS (Tris-buffered saline) for 15 minutes, and washed with TBS (two times, 5 minutes each). The slides were blocked using 5% donkey serum in TBSS (TBS with 0.1% saponin) for 60 minutes at room temperature and then incubated overnight at 4˚C with primary antibody (Rabbit anti- SCGB1A1, 1:200). The slides were washed three times (5 minutes each) with TBSS and incubated with secondary antibody at 1:500 for 60 minutes at room temperature. The slides were washed two times (5 minutes each) with TBSS and mounted using DAPI Fluoromount-G (Cat#0100-20, Southern Biotech). For quantification of both SCGB1A1 and MUC5B the above protocol was followed with the following changes: antigen retrieval was performed using 10mM Sodium Citrate buffer (pH 9.0) with 0.05% Tween-20, slides were blocked using 5% goat serum in TBSS as above and then incubated overnight at $4^{o}$C with primary antibody (Mouse anti- SCGB1A1, 1:100; Rabbit anti-MUC5B, 1:100; or both). Fluorescent images were taken using a Leica widefield fluorescence microscope.

Automated quantification of immunofluorescence imaging was performed using *ImageJ*. Monochrome images were thresholded to ensure removal of background autofluorescence. The percent of SCGB1A1-stained cells was quantified using the *Analyze Particles* command by setting the size threshold to 1 $\mu m^2$ and using all other default settings. Counting DAPI+ cells was performed in a similar manner. For each slide, the ratio of SCGB1A1+ particle area to DAPI+ particle number was calculated for each of 42 images (20X magnification) from each control and COVID-19 lung sample. One-tenth of the images with SCGB1A1+ cells from each sample were randomly selected and analyzed by manually counting the number of SCGB1A1+ airway epithelial cells in airways measuring > or <100 µm in diameter. Manual counting the number of SCGB1A1+, MUC5B+, and dual SCGB1A1+/MUC5B+ airway epithelial cells in COVID-19 explants and sex-matched controls (n=7 each) were performed using 5x5 tile scans.

**Statistical Analyses**

In analyses described in Figure 1, we adjusted for a false discovery rate of 0.05 based on biomarker class and category of association using the Benjamini-Hochberg method.[1] To ensure that our results were robust to potential model overfitting, we examined all GAM plots and conducted sensitivity analyses excluding visible outliers >2 standard deviations that persisted after natural log transformation. Analyses were performed using Stata v16 (StataCorp) and R studio (v2024.04.2+764).

**Materials**

Antibodies include Rabbit anti-SCGB1A1 (Cat#10490-1-AP, Proteintech); Mouse anti-SCGB1A1 (Cat# sc-365992, Santa Cruz), Rabbit anti-MUC5B (Cat# HPA008246; Sigma-Aldrich), Donkey anti-rabbit IgG (H+L) Alexa Fluor Plus 647 (Cat#A32795, Invitrogen), Goat anti-mouse IgG (H+L) Alexa Fluor 488 (Cat#A11001, Invitrogen).

**Figure E1. Hematoxylin and eosin staining of transbronchial biopsies from four asymptomatic lung transplant recipients done as part of routine surveillance during the first year after lung transplantation.** Since all biopsies showed preserved lung alveolar architecture without evidence of infection or rejection, these samples were used as healthy control lung tissue samples in single-cell analyses. Demographics of these control subjects are described in Table E8.

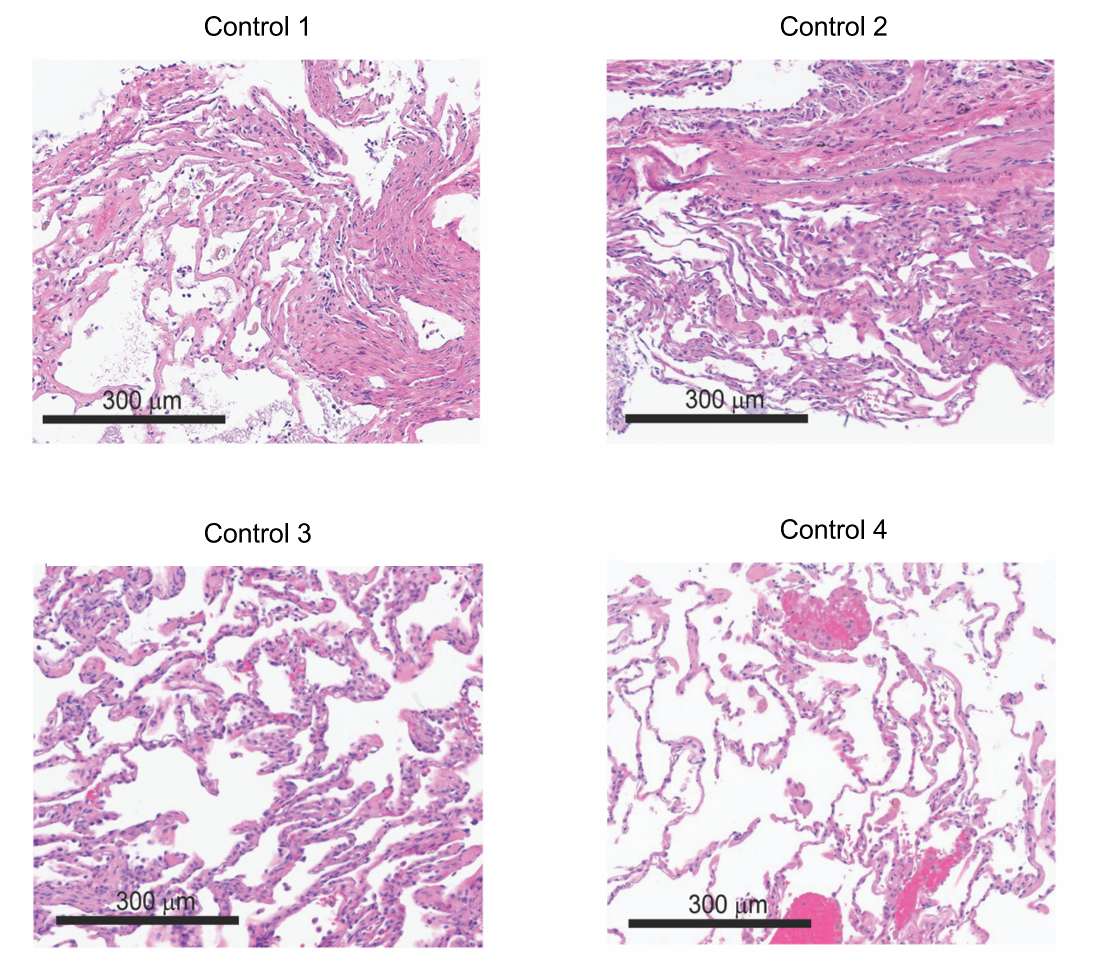

**Figure E2**. Columbia participant follow-up visit participation.

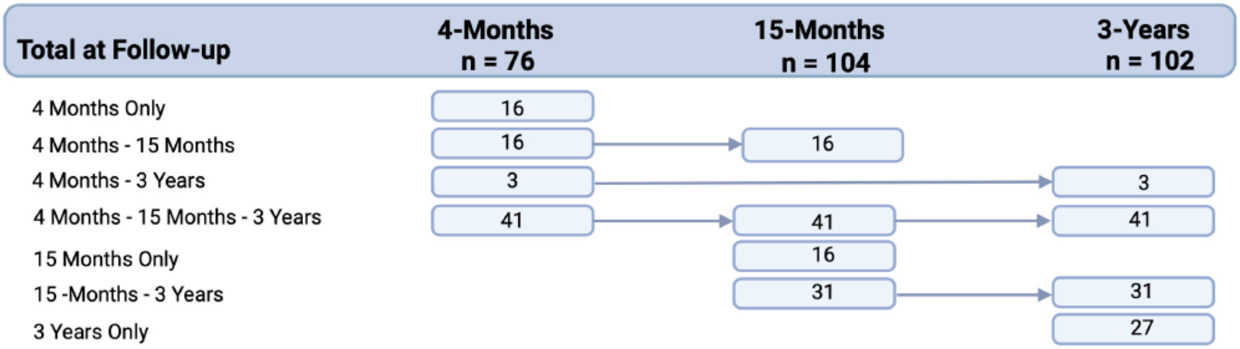

**Table E1**. Demographic and clinical characteristics stratified by follow-up visit in the Columbia (discovery) cohort.

| **Characteristics** | **Columbia 4-month follow-up (n=76)** | **Columbia 15-month follow-up (n=104)** | **Columbia 3-year follow-up (n = 102)** |
|---|---|---|---|
| Age, years, mean (SD) | 54 (14) | 54 (11) | 56 (12) |
| Male | 45 (61%) | 61 (59%) | 55 (54%) |
| Body Mass Index (kg/m$^2$), mean (SD) | 32 (6.9) | 32 (6.8) | 32 (6.2) |
| Hispanic, n (%) | 43 (57%) | 62 (60%) | 69 (68%) |
| Race, n (%) | | | |
| White | 30 (39%) | 30 (29%) | 40 (40%) |
| Black | 22 (30%) | 28 (27%) | 25 (25%) |
| Asian | 1 (1%) | 4 (4%) | 4 (4%) |
| Other | 23 (30%) | 42 (40%) | 33 (32%) |
| **Comorbidities** | | | |
| Ever smoker* | 31 (41%) | 40 (38%) | 39 (38%) |
| Active smoker | 2 (3%) | 3 (3%) | 2 (2%) |
| COPD | 4 (5%) | 4 (4%) | 2 (2%0 |
| Asthma | 18 (27%) | 24 (23%) | 22 (22%) |
| Cardiovascular disease | 7 (9%) | 13 (13%) | 13 (13%) |
| eGFR, mean (SD) | 96 (44) | 98 (45) | 98 (45) |
| **Acute COVID-19 Illness Characteristics** | | | |
| Admission SOFA score, median [IQR] | 3.5 [3-6] | 3 [2-5] | 3 [2-5] |
| Steroid use, n (%) | 39 (51%) | 43 (41%) | 47 (46%) |
| IL-6 receptor inhibitor use, n (%) | 17 (22%) | 26 (25%) | 23 (23%) |
| Oxygen use | | | |
| None | 0 (0%) | 0 (0%) | 0 (0%) |
| Nasal Cannula | 23 (30%) | 31 (30%) | 32 (31%) |
| Non-Rebreather | 17 (22%) | 18 (17%) | 18 (18%) |
| NIPPV or HFNO | 4 (5%) | 2 (2%) | 2 (2.0%) |
| Mechanical ventilation, n( %) | 32 (42%) | 53 (51%) | 50 (49%) |
| Ventilator days, median (IQR) | 31 [12-42] | 35 [18-46] | 35 [17-46] |
| Hospital Days, median [IQR] | 18 [7-35] | 23 [8-47] | 23 [7-46] |

*Ever smoker: former or active smoker. COPD: chronic obstructive pulmonary disease. eGFR: estimated glomerular filtration rate based upon CKD-EPI equation.[2] SOFA: sequential organ failure assessment. NIPPV: non-invasive positive pressure ventilation. HFNO: high-flow nasal oxygen.

**Table E2**. Fibrotic-like abnormalities on Thoracic CT scans in the Columbia (discovery), University of British Columbia (validation), and McGill (validation) cohorts.

| **Cohort** | **Columbia 4-month follow-up (n=76)** | **Columbia 15-month follow-up (n=104)** | **Columbia 3-year follow-up (n = 102)** | **UBC 3-month follow-up (n = 56)** | **McGill Hospital DC, 4-month, or 15-month follow-up (n=37)** |
|---|---|---|---|---|---|
| Months from admission to thoracic CT, median [IQR] | 4.5 [4.0-4.8] | 15 [14.2-16.5] | 39 [37-42] | 3.2 [2.4-3.7] | 2.6 [0.13-5.7] |
| Any fibrotic pattern, n (%) | 45 (59%) | 67 (64%) | 62 (62%) | 32 (57%) | 16 (43%) |
| Reticulations, n (%) | 30 (39%) | 67 (64%) | 58 (57%) | 31 (55%) | 15 (41%) |
| Traction Bronchiectasis, n (%) | 21 (28%) | 41 (39%) | 46 (45%) | 28 (50%) | 5 (14%) |
| Honeycombing, n (%) | 1 (1.0%) | 1 (1.0%) | 1 (1.0%) | 2 (3.6%) | 4 (11%) |

**Table E3**. Comparison of demographic and clinical characteristics between those with and without fibrotic-like abnormalities in the McGill (validation) nested case-control cohort.

| | **Case: Fibrotic-Like Abnormalities (n=16)** | **Control: No Fibrotic-Like Abnormalities (n = 21)** | **p-value** |
|---|---|---|---|
| Age, mean (SD) | 67 (10) | 63 (18) | 0.36 |
| Male, n (%) | 12 (75%) | 10 (48%) | 0.18 |
| Body Mass Index (kg/m2), mean (SD) | 28 (4.0) | 30 (6.6) | 0.34 |
| Race/Ethnicity, n (%) | | | |
| White | 11 (69%) | 13 (62%) | 0.72 |
| Asian | 2 (13%) | 2 (10%) | |
| Hispanic | 2 (13%) | 1 (4.8%) | |
| Middle Eastern | 1 (6.2%) | 3 (14%) | |
| Other | 0 (0%) | 1 (4.8%) | |
| **Comorbidities** | | | |
| Smoking status, n (%) | | | |
| Never | 9 (56%) | 16 (76%) | 0.17 |
| Active | 0 (0%) | 1 (4.8%) | |
| Ever | 7 (44%) | 3 (14%) | |
| Unknown | 0 (0%) | 1 (4.8%) | |
| COPD, n (%) | 6.2 (1%) | 0 (0%) | 0.89 |
| Asthma, n (%) | 2 (13%) | 0 (0%) | 0.35 |
| Cardiovascular disease, n (%) | 5 (31%) | 4 (19%) | 0.64 |
| eGFR, mean (SD) | 82.3 (24.2) | 83.4 (27.6) | 0.9 |
| **Acute COVID-19 Illness Characteristics** | | | |
| Corticosteroid Use, n (%) | 12 (75%) | 11 (52%) | 0.29 |
| IL-6 receptor inhibitor use, n (%) | 1 (6.2%) | 0 (0%) | 0.89 |
| Oxygen Use, n (%) | | | 0.83 |
| None | 4 (25%) | 3 (14%) | |
| Nasal Cannula | 10 (62%) | 16 (76%) | |
| NIPPV or HFNO | 1 (6.2%) | 1 (4.8%) | |
| Mechanical Ventilation | 1 (6.2%) | 1 (4.8%) | |
| Ventilator days, median [IQR] | 12 [10-13] | 33 [33-33] | 0.22 |
| Hospital days, median [IQR] | 9 [5.8-20] | 6 [4.0-16] | 0.43 |
| Days from admission to CT scan, median [IQR] | 99 [32-147] | 8 [0-425] | 0.19 |

*Ever smoker: former or active smoker. COPD: chronic obstructive pulmonary disease. eGFR: estimated glomerular filtration rate based on CKD-EPI equation.[2] NIPPV: non-invasive positive pressure ventilation. HFNO: high-flow nasal oxygen.

**Table E4**. Effect estimates of associations between CC16 and fibrotic-like lung abnormalities on thoracic CT scan in the Columbia (discovery) cohort.

| | CC16 at hospital discharge | | |
|---|---|---|---|
| | **Fibrotic abnormalities at 4 Months** | **Fibrotic abnormalities at 15 Months** | **Fibrotic abnormalities at 3 Years** |
| Effect per tertile increase in CC16 (pg/ml) | OR (95% CI) | OR (95% CI) | OR (95% CI) |
| Low | 1 | 1 | 1 |
| Medium | 2.27 (0.47, 13.1) | 1.91 (0.56, 7.0) | 9.32 (1.97, 56.2) |
| High | 10.0 (2.12, 62.44) | 14.3 (2.92, 112) | 21.2 (3.76, 188) |
| p-value for trend | 0.007 | 0.002 | 0.001 |
| Effect per 1 natural log-fold increase in CC16 (pg/ml) | 4.36 (1.43, 17.4) | 10.75 (3.02, 55.1) | 23.88 (4.20, 280) |
| p-value | 0.02 | 0.001 | 0.003 |

| | CC16 at 4-months | | |
|---|---|---|---|
| | **Fibrotic abnormalities at 4 Months** | **Fibrotic abnormalities at 15 Months** | **Fibrotic abnormalities at 3 Years** |
| Effect per tertile increase in CC16 (pg/ml) | OR (95% CI) | OR (95% CI) | OR (95% CI) |
| Low | 1 | 1 | 1 |
| Medium | 2.03 (0.55, 8.08) | 5.97 (1.43, 29.3) | 3.27 (0.65, 18.7) |
| High | 10.0 (2.62, 45.6) | 16.8 (3.20, 137) | 21.2 (2.75, 465) |
| p-value for trend | 0.002 | 0.002 | 0.007 |
| Effect per 1 natural log-fold increase in CC16 (pg/ml) | 7.39 (2.52, 27.4) | 17.0 (4.09, 112) | 8.64 (2.17, 50.6) |
| p-value | 0.0009 | 0.0006 | 0.006 |

| | CC16 at 15-months | | CC16 at 3-years |
|---|---|---|---|
| | **Fibrotic abnormalities at 4 Months** | **Fibrotic abnormalities at 15 Months** | **Fibrotic abnormalities at 3 Years** |
| Effect per tertile increase in CC16 (pg/ml) | OR (95% CI) | OR (95% CI) | OR (95% CI) |
| Low | 1 | 1 | 1 |
| Medium | 2.65 (0.90, 8.16) | 2.15 (0.63, 7.65) | 3.02 (1.12, 8.54) |
| High | 9.38 (2.80, 38.4) | 6.83 (1.76, 31.8) | 10.19 (3.27, 37.3) |
| p-value for trend | 0.0005 | 0.008 | 0.0001 |
| Effect per 1 natural log-fold increase in CC16 (pg/ml) | 5.81 (2.10, 19.6) | 5.04 (1.55, 20.7) | 6.57 (2.67, 19.0) |
| p-value | 0.002 | 0.014 | 0.0002 |

**Table E5**. Cross-sectional spearman correlations of CC16 and percent predicted diffusion capacity of lung for carbon monoxide (DLCO) at follow-up in the Columbia (discovery) and University of British Columbia (validation) cohorts.

| Columbia | Rho | p-value |
|---|---|---|
| 4 Months | -0.54 | <0.001 |
| 15 Months | 0.01 | 0.92 |
| 3 years | -0.33 | <0.001 |
| UBC | | |
| 3 months | -0.39 | 0.004 |

**Table E6**. Effect estimates of associations between CC16 and fibrotic-like lung abnormalities on chest CT scan in the validation cohorts.

| | |
|---|---|
| | **University of British Columbia (n = 56)** |
| | **Adjusted association of CC16 with Fibrotic abnormalities at 3 Months** |
| Effect per tertile increase in CC16 (pg/ml) | OR (95% CI) |
| Low | 1 |
| Medium | 1.89 (0.53 - 7.09) |
| High | 3.56 (0.93 - 15.2) |
| p-value for trend | 0.0763 |
| Effect per 1 natural log-fold increase in CC16 (pg/ml) | 2.92 (1.07 - 9.07) |
| p-value | 0.0462 |
| | **McGill University (n = 37)** |
| | **Adjusted association of CC16 at hospital discharge with fibrotic abnormalities at hospital discharge, 4 months, or 15 months** |
| Effect per tertile increase in CC16 (pg/ml) | |
| Low | 1 |
| Medium | 1.43 (0.24, 9.05) |
| High | 5.41 (0.79, 52.5) |
| p-value for trend | 0.11 |
| Effect per 1 natural log-fold increase in CC16 (pg/ml) | 2.36 (0.93, 7.03) |
| p-value | 0.088 |
| | **McGill University Sensitivity Analysis (n = 34)** |
| | **Adjusted association of CC16 at hospital discharge with fibrotic abnormalities at hospital discharge, 4 months, or 15 months** |
| Effect per tertile increase in CC16 (pg/ml) | |
| Low | 1 |
| Medium | 2.02 (0.24, 19.1) |
| High | 3.37 (0.48, 28.9) |
| p-value for trend | 0.23 |
| Effect per 1 natural log-fold increase in CC16 (pg/ml) | 2.11 (0.79, 6.67) |
| p-value | 0.158 |

Associations are adjusted for age, sex, race/ethnicity, body mass index, smoking history, COPD, asthma, estimated glomerular filtration rate, use of corticosteroids, IL-6 receptor inhibitor therapy, ventilator days during the hospitalization for acute COVID-19, and days between SARS-CoV2 infection and thoracic CT scan.

**Figure E3.** Circulating CC16 (pg/ml) levels in those with and without fibrotic-like abnormalities in the McGill (validation) nested case-control cohort, stratified by follow-up time period of thoracic CT scan: (A) hospital discharge, (B) 4 months; (C) 15 months. Sensitivity analysis

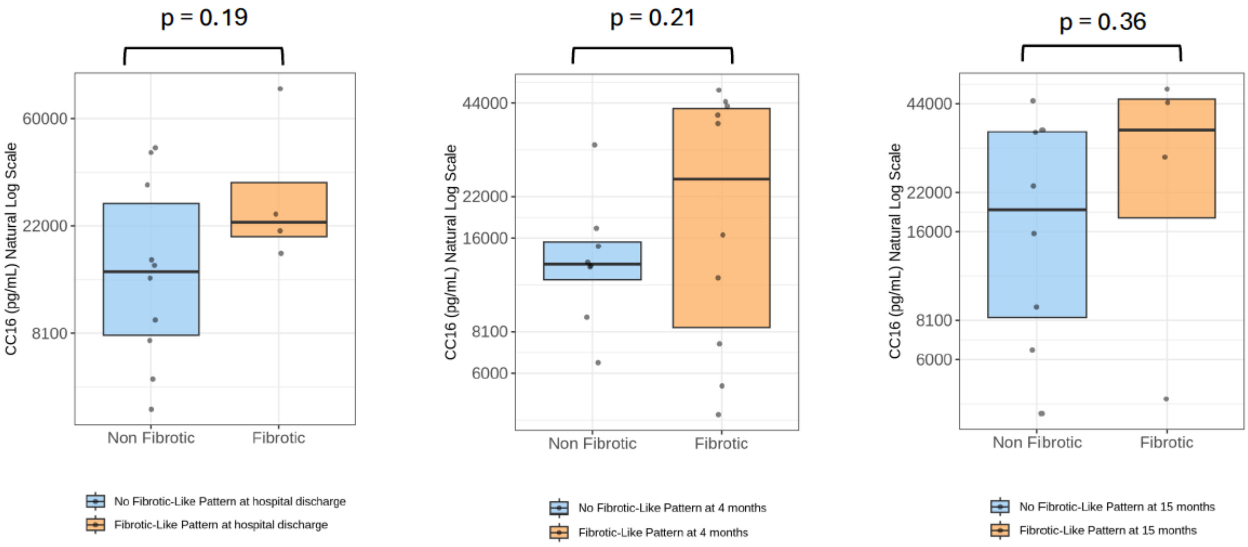

**Figure E4.** Circulating CC16 (pg/ml) levels in those with and without fibrotic-like abnormalities in the McGill (validation) nested case-control cohort sensitivity analysis excluding n=3 case patients with honeycombing in the McGill cohort (A), and stratified by follow-up time period of thoracic CT scan: (B) hospital discharge, (C) 4 months; (D) 15 months. Sensitivity analysis. Generalized additive model (GAM) with LOESS smoothers with adjustment for age, sex, race/ethnicity, BMI, COPD, pack-year history of smoking, estimated glomerular filtration rate, ventilator days, and days since initial SARS-CoV-2 infection using covariate balanced propensity scores.

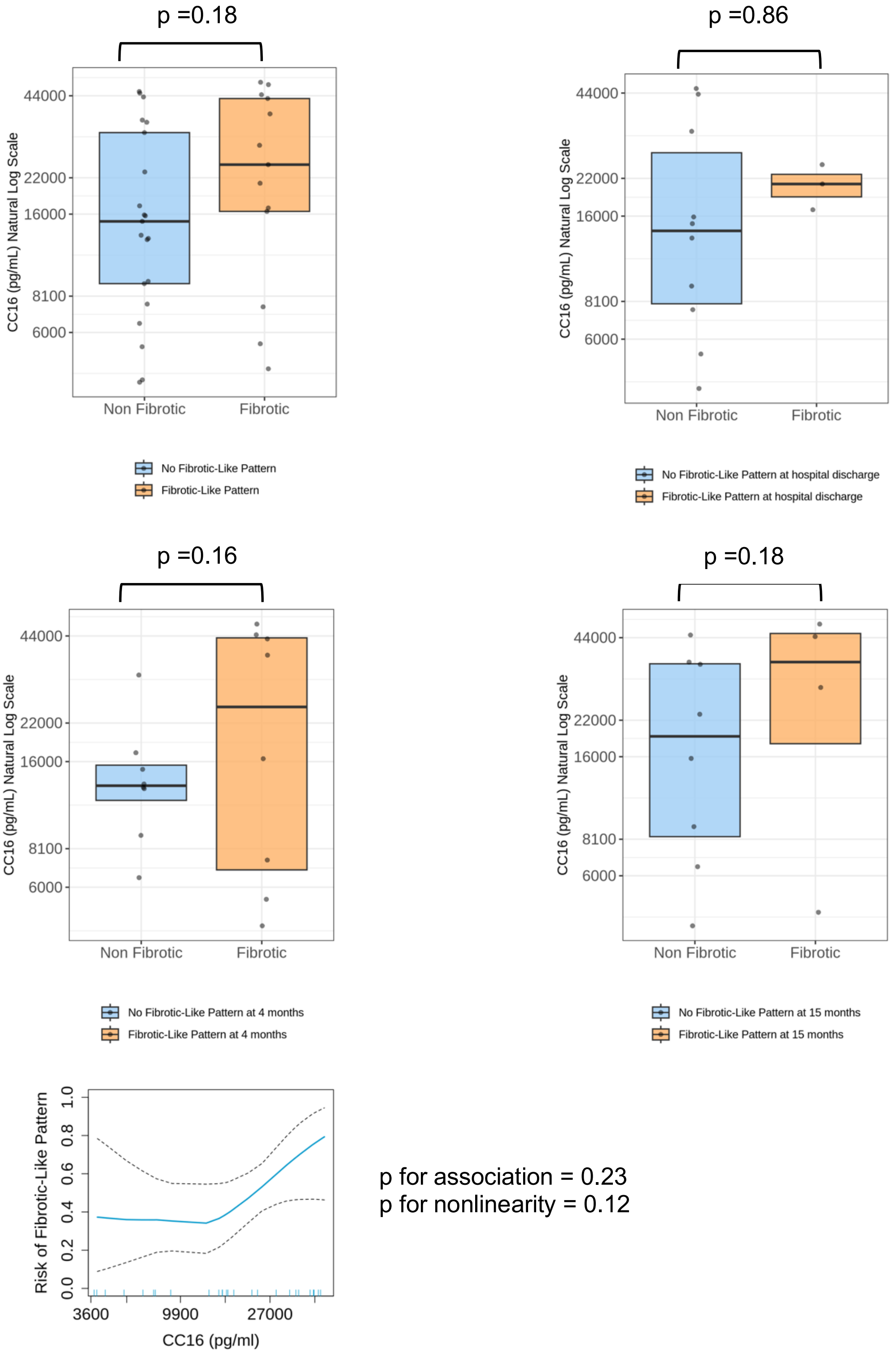

**Table E7**. Generalized linear model adjusted associations of CC16 with airway-to-lung ratio 15 months in Columbia (discovery) cohort.

| **Association** | **unit change in airway-to-lung ratio per natural log-fold increase in CC16, β (95% CI)** | **SD-unit change in A/L ratio per natural-log fold increase in CC16, β (95% CI)** | **p-value** |
|---|---|---|---|
| Hospital Discharge CC16 - 15-month thoracic CT scan | 0.30 (0.10, 0.49) | 0.61 (0.21, 1.01) | 0.003 |
| 4-Month CC16 - 15-month thoracic CT scan | 0.29 (0.07, 0.45) | 0.58 (0.15, 1.02) | 0.01 |
| 15-month CC16 - 15-month thoracic CT scan | 0.36 (0.18, 0.54) | 0.72 (0.36, 1.09) | <0.001 |

Associations are adjusted for age, sex, race/ethnicity, body mass index, smoking history, COPD, asthma, estimated glomerular filtration rate, use of corticosteroids, IL-6 receptor inhibitor therapy, ventilator days during the hospitalization for acute COVID-19, and days between SARS-CoV2 infection and thoracic CT scan

**Figure E5**. Sensitivity analyses of generalized additive model adjusted associations between CC16 and airway to lung ratio. (A-C) Complete case analyses among those with all subsegmental airways visualized. (D-F) Analyses of all subjects excluded sub-segmental measurements.

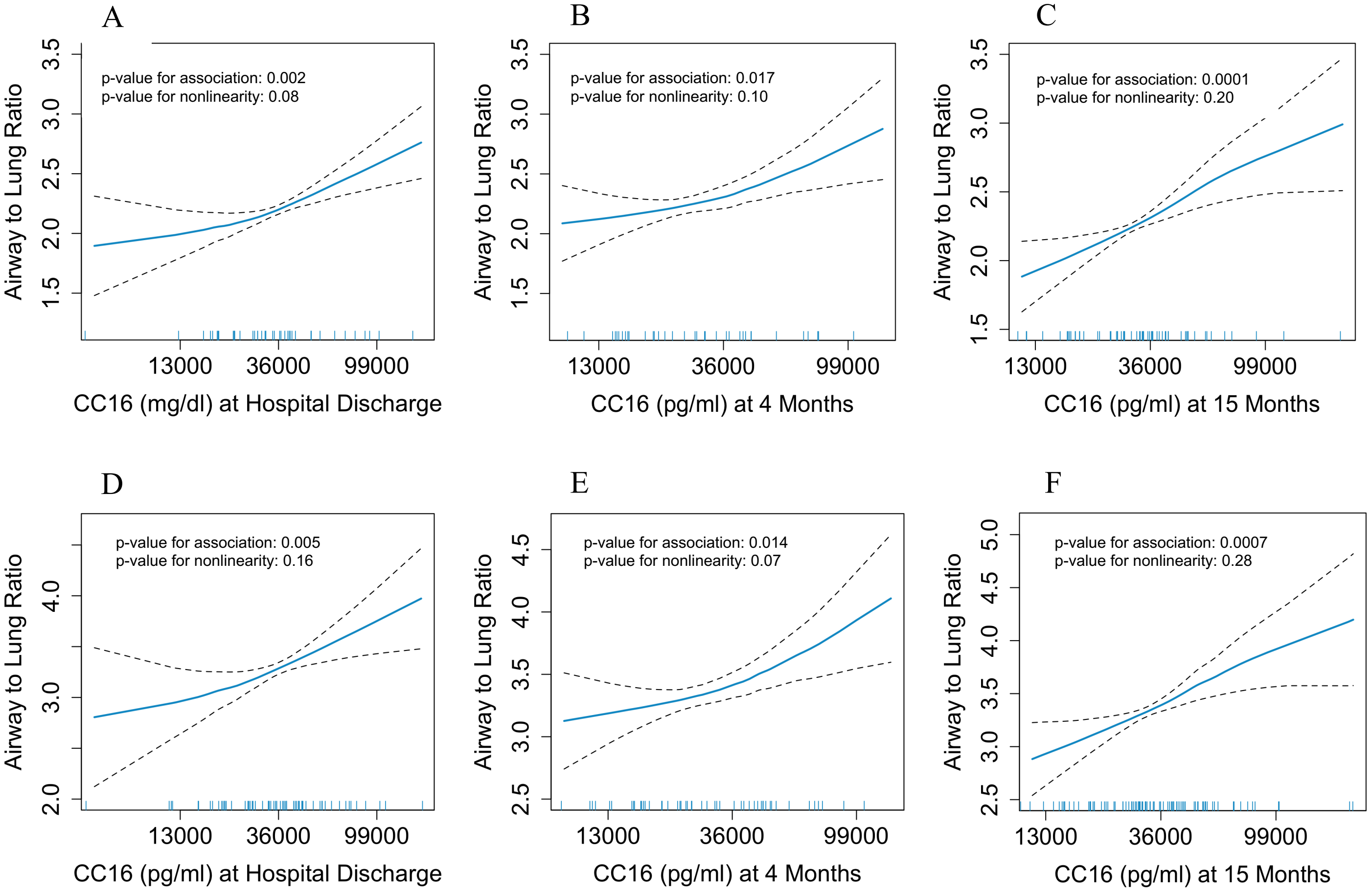

**Table E8.** Demographics of COVID-19 pulmonary fibrosis and control patients who underwent bronchoscopy.

| Group | Age, years | Sex | Race/Ethnicity | Time in months from COVID-19 infection until bronchscopy | Time in months from lung transplant until bronchoscopy (indication for transplant) |
|---|---|---|---|---|---|
| COVID-19 | 56-60 | Male | Hispanic | 50 | n/a |
| COVID-19 | 36-40 | Male | Hispanic | 50 | n/a |
| COVID-19 | 61-65 | Female | Hispanic | 52 | n/a |
| COVID-19 | 56-60 | Male | Hispanic | 51 | n/a |
| Control | 36-40 | Female | White | n/a | 9 mo (BOS) |
| Control | 66-70 | Female | Asian | n/a | 9 mo (non-CF bronch.) |
| Control | 46-50 | Male | White | n/a | 3 mo (sarcoidosis) |
| Control | 61-65 | Male | White | n/a | 11 mo (COPD/emphysema) |

n/a: not applicable. BOS, bronchiolitis obliterans syndrome, non-CF bronch., non-cystic fibrosis bronchiectasis; COPD, chronic obstructive pulmonary disease

**Figure E6. Single cell RNA sequencing quality control, gene expression data, and differences in cell numbers of immune, endothelial, and non-epithelial, non-endothelial, and non-immune cells. A**. Proportion of EpCAM+ (blue), CD31+CD45+ (red), and negative (green) cells as determined by expression of *EPCAM*, *PTPRC* (CD45), *PECAM1* (CD31), or none of these genes. One COVID-19 sample which had a low viability and a high proportion of EpCAM-CD31-CD45- cells was not included in the study, leaving a total of four COVID-19 biopsies being used in analyses. Three lung transplant participants had evidence of rejection, and were therefore excluded from the study, leaving a total of 4 lung transplant recipient biopsies being used as controls. A total of eight samples (n = 4 COVID explants and n = 4 lung transplant controls) were included in the single cell RNA sequencing analyses. A total of 41,003 post-COVID cells and 23,516 control cells were included in the analyses. **B**. Feature plot of epithelial cell clusters identified in this study and the genes that characterize each cell cluster **C.** Proportion of different subsets of lung immune and endothelial (CD13+CD45+) cells found in control and COVID-19 samples. **D.** Proportion of different subsets of lung non-epithelial, non-endothelial, and non-immune (EpCAM-CD13-CD45-) cells found in control and COVID-19 samples.

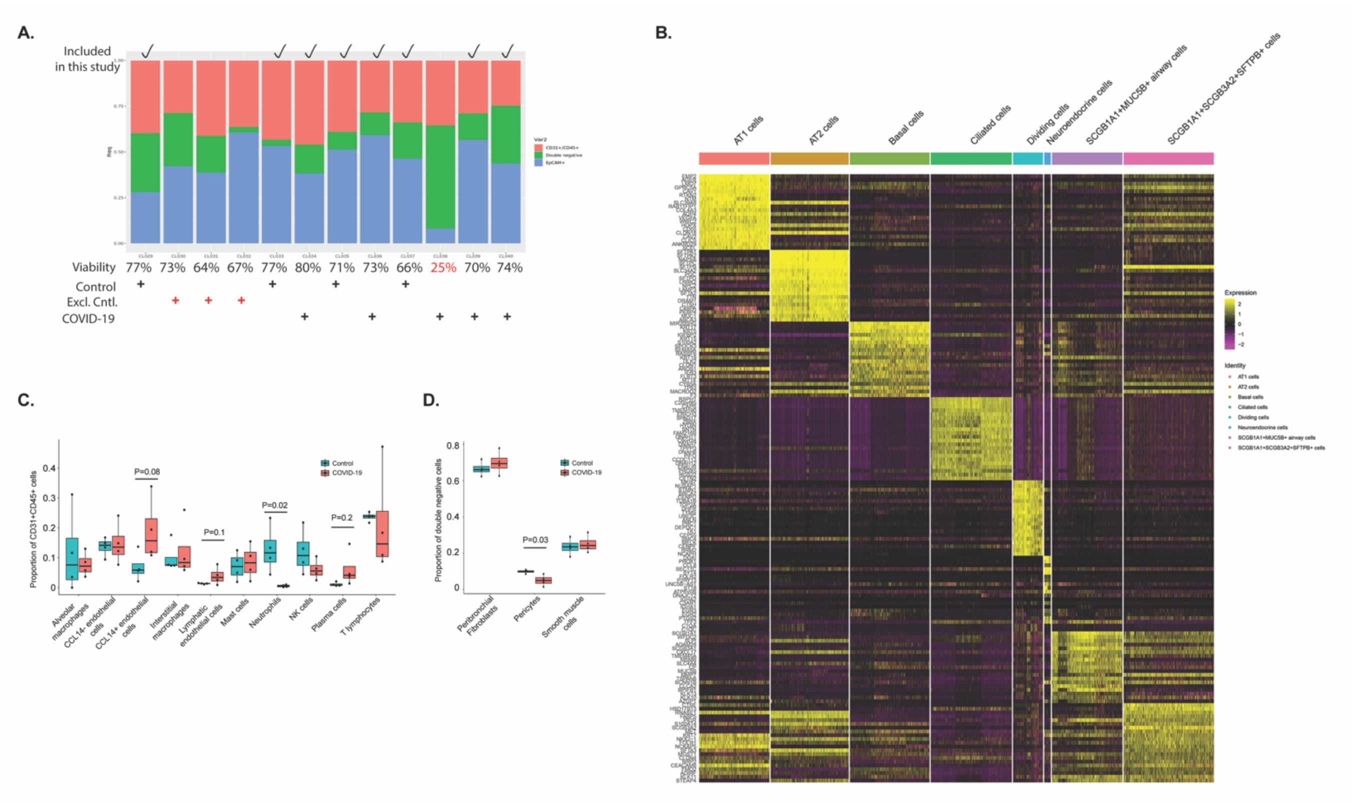

**Table E9.** Demographics of patients who underwent lung transplantation for COVID-19 pulmonary fibrosis and controls who underwent surgical resection of pulmonary nodules.

| **Group** | **Age, years** | **Sex** | **Race/Ethnicity** | **Time in months from COVID-19 infection until lung transplant** |
|---|---|---|---|---|
| COVID-19 | 61-65 | Male | White | 3 |
| COVID-19 | 56-60 | Male | White | 3 |
| COVID-19 | 61-65 | Male | Hispanic | 12 |
| COVID-19 | 51-55 | Female | Hispanic | 2 |
| COVID-19 | 46-50 | Female | Unknown | 7 |
| COVID-19 | 61-65 | Female | Hispanic | 6 |
| COVID-19 | 66-70 | Male | Hispanic | 29 |
| Control | 46-50 | Female | White | n/a |
| Control | 46-50 | Female | White | n/a |
| Control | 76-80 | Female | Black | n/a |
| Control | 51-55 | Female | Black | n/a |
| Control | 61-65 | Female | Hispanic | n/a |
| Control | 71-75 | Female | White | n/a |
| Control | 66-70 | Female | White | n/a |
| Control | 66-70 | Male | White | n/a |
| Control | 51-55 | Male | White | n/a |
| Control | 66-70 | Male | White | n/a |
| Control | 61-65 | Male | Other | n/a |

n/a: not applicable.

**E-REFERENCES**

1. Benjamini Y. Controlling the False Discovery Rate: A Practical and Powerful Approach to Multiple Testing. *Journal of the Royal Statistical Society Series B: Statistical Methodology*; **57**(1): 289-300.

2. Delgado C, Baweja M, Crews DA-O, et al. A Unifying Approach for GFR Estimation: Recommendations of the NKF-ASN Task Force on Reassessing the Inclusion of Race in Diagnosing Kidney Disease. (1533-3450 (Electronic)).
